# Mapping Epigenetic Gene Variant Dynamics: Comparative Analysis of Frequency, Functional Impact and Trait Associations in African and European Populations

**DOI:** 10.1101/2024.08.11.24311816

**Authors:** Musalula Sinkala, Gaone Retshabile, Phelelani T. Mpangase, Salia Bamba, Modibo K Goita, Vicky Nembaware, Samar S. M. Elsheikh, Jeannine Heckmann, Kevin Esoh, Mogomotsi Matshaba, Clement A. Adebamowo, Sally N. Adebamowo, Ofon Elvis Amih, Ambroise Wonkam, Michele Ramsay, Nicola Mulder

## Abstract

Epigenetic modifications influence gene expression levels, impact organismal traits, and play a role in the development of diseases. Therefore, variants in genes involved in epigenetic processes are likely to be important in disease susceptibility, and the frequency of variants may vary between populations with African and European ancestries. Here, we analyse an integrated dataset to define the frequencies, associated traits, and functional impact of epigenetic gene variants among individuals of African and European ancestry represented in the UK Biobank. We find that the frequencies of 88.4% of epigenetic gene variants significantly differ between these groups. Furthermore, we find that the variants are associated with many traits and diseases, and some of these associations may be population-specific owing to allele frequency differences. Additionally, we observe that variants associated with traits are significantly enriched for quantitative trait loci that affect DNA methylation, chromatin accessibility, and gene expression. We find that methylation quantitative trait loci account for 71.2% of the variants influencing gene expression. Moreover, variants linked to biomarker traits exhibit high correlation. We therefore conclude that epigenetic gene variants associated with traits tend to differ in their allele frequencies among African and European populations and are enriched for QTLs.

## Introduction

Epigenetic variations that result in changes in gene expression levels are important in determining traits and diseases. Consequently, single nucleotide polymorphisms (SNPs) in genes involved in epigenetic mechanisms (hereafter referred to as epigenetic genes for simplicity) might therefore be important in disease susceptibility^1,2^. Furthermore, epigenetic processes such as DNA methylation, chromatin remodelling, and covalent histone modifications can be just as crucial in traits and diseases ^3,4^. Expectedly, SNPs in regions encoding epigenetic genes are associated with various traits and diseases ^3,5–7^.

An integrative analysis of 111 reference human epigenomes revealed that disease- and trait-associated genetic variants are enriched in tissue-specific epigenomic markers, emphasizing the relevance of epigenomic information in understanding gene regulation, cellular differentiation, and human disease ^8^. Other studies have revealed widespread genetic variations affecting the regulation of most genes, illuminating the cellular mechanisms of transcriptome variations and the landscape of functional variants in the human genome ^9^. Notably, most previous studies have applied these analyses primarily to European (EUR) ancestry populations ^10,11^.

There are documented cases of SNPs that vary significantly among populations of different genetic ancestry backgrounds ^8,9,12,13^. For instance, African (AFR) populations are known to have a more variable genome than other populations ^14–16^. A study examining worldwide patterns of human epigenetic variations reported that population-specific DNA methylation, an important epigenetic mechanism, mirrors genomic variations and exhibits greater local genetic control than mRNA levels, showing the importance of epigenetic factors in genetic diversity across different populations ^17^.

Determining the interplay between genetic variability (SNPs) and epigenetic variations should allow us to uncover associated traits and diseases. In the past decade, large-scale genome sequencing has deepened our insights into genetic variations, highlighting the diversity within AFR populations compared to their more extensively studied EUR counterparts. For example, recent genome-wide association studies (GWAS) identified several genetic loci associated with different traits with differences in the SNP allele frequencies between AFRs and EURs ^11,18–20^.

The UK Biobank (UKB) cohort contains data on 502,462 individuals, providing an opportunity to compare the frequency of genomic variants among different populations and associations with traits among individuals of EUR and recent AFR descent ^21^. By integrating allele frequencies, genetic associations of traits, and the functional impact of SNPs, it is possible to identify population-specific loci, underscoring the importance of considering both genetic uniqueness and shared genetic factors in AFRs and EURs ^11,22,23^. Identifying population-specific epigenetic gene variants associated with traits will provide additional information relevant to understanding various traits and diseases in distinct populations ^24–26^. However, to our knowledge, there is no information on the variations in epigenetic gene SNP frequencies across the entire UKB cohort or separately among AFRs and EURs.

Here, we compare the allele frequencies of SNPs in epigenetic genes and traits associated with variants at epigenetic gene loci among individuals of AFR and EUR ancestry represented in the UKB and other AFR datasets, utilising genotype array data with imputation to enhance variant detection. First, we consolidate data from multiple databases to pinpoint trait associations with variants in epigenetic genes, as identified in published GWAS. This was followed by analyses to define the functional impact of the epigenetic variants. Furthermore, we evaluated the variant frequencies and association with several biomarker levels among UKB-AFR and UKB-EUR individuals. This approach allowed us to identify epigenetic loci associated with various traits among UKB-AFRs and UKB-EURs, with some loci showing a high likelihood of modifying these traits in a population-specific manner.

## Results

We analysed an imputed UKB^21^ SNP genotype array dataset with individual-level phenotypic and clinical characteristics for 383,471 EUR (UKB-EUR) and 5,978 individuals of recent AFR (UKB-AFR) descent. In our study, ancestries were determined through a combination of self-identification by participants in the UKB dataset, followed by refinement using principal component analysis (PCA) and a random forest algorithm based on PCA results, enabling accurate reassignment to self-identified ancestries with a membership posterior probability above 0.5, in line with UKB standards ^21^.

Furthermore, we curated a list of 283 epigenetic genes (genes involved in epigenetic mechanisms) and epigenetic regulator genes from the Reactome pathways knowledgebase^27,28^ and the literature^29^. Among these 283 epigenetic genes, the largest number encodes histone methyltransferases (HMT; 62 genes), the genes containing a bromodomain (Bromo; 35 genes), and histone deacylases (HDAC; 33 genes) (Figure 1a, Supplementary File 1).

**Figure 1:**
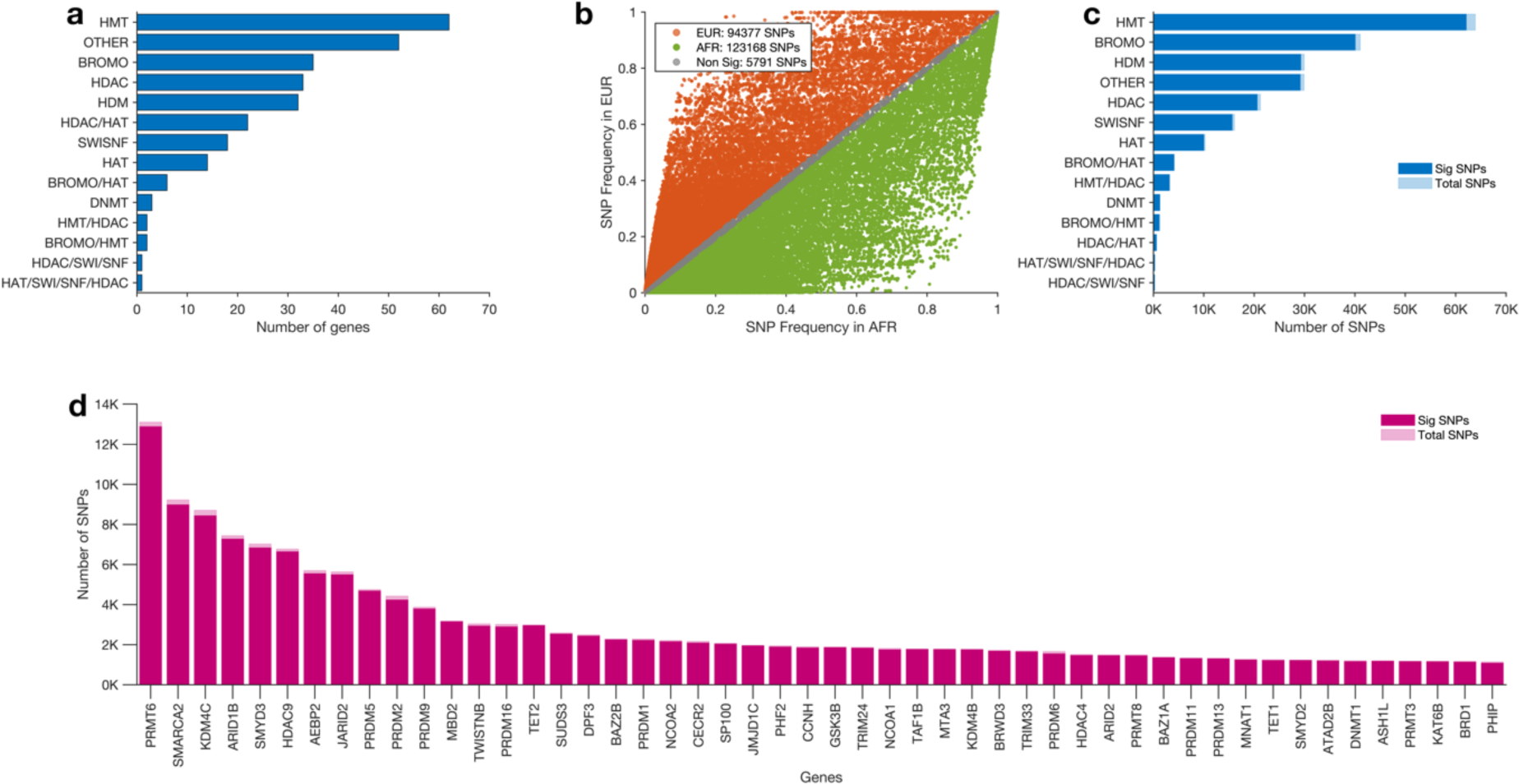
Epigenetic genes and gene variants. (a) Distribution of 283 epigenetic genes across 14 classes of epigenetic genes. (b) scatter plot showing the frequency of SNPs in epigenetic genes among UKB-AFR and UKB-EUR. The colours show details about statistical significance: orange for variants significantly more frequent in UKB-EUR, green for variants significantly more frequent in AFR, and grey for variants whose frequencies are similar in UKB-AFR and UKB-EUR. (c) The number of variants found in each class of epigenetic genes. (d) Number of variants in the top 20 epigenetic genes with the most variants.

### The frequency of SNPs varies between AFRs and EURs

Next, we extracted a list of 223,336 high-confidence variants, each with a frequency greater than 1 x 10^-5^, at loci "near/within" epigenetic genes. As detailed in the Methods section, these loci span regions up to 500 kb upstream or downstream. We then compared the minor allele frequencies of these variants between UKB-AFR and UKB-EUR populations. We observed, through Fisher’s exact test, that 123,168 (55.1%) of the 223,336 variant alleles were significantly more frequent in AFRs (Benjamini-Hochberg adjusted p-values < 0.05), 94,377 (42.3%) were more frequent in UKB-EURs, and 5,791 (2.6%) were distributed similarly in the two populations (See Figure 1b and Supplementary Figures 1a to 1c). To ensure a comprehensive exploration of genetic diversity across populations, we did not apply a frequency cut-off point in our analysis, allowing for an inclusive examination of rare and common variants in line with previous studies ^30,31^.

We found that the HMT gene class exhibited the highest number of variants, totalling 63,896, of which 62,088 (97.2%) were significantly different in frequency between UKB-EUR and UKB-AFR individuals. Nearby or within genes encoded by the BROMO class, we identified 41,065 variants, with 40,048 (97.5%) showing significant differences between UKB-AFR and UKB-EUR populations. For the OTHER category, we identified 29,903 variants, with 29,141 (97.5%) exhibiting significant frequency differences (see Figure 1c). The HMT gene class had the most variants, even after adjusting for gene length within each class (Supplementary Figure 2a).

Furthermore, we found the highest number of variants (13,098) at the loci near the *PRMT6* gene within the HMT class, of which 12,877 (98.3%) were significantly different. SMARCA2 followed this with 9,223 variants, of which 8,979 (97.4%) were significantly different, and *KDM4C* with 8,705 variants, of which 8,433 (96.9%) were significantly different (see Figure 1d). We found the most variants at the loci within or near the *PRMT6* gene, even after normalising the number of variants to the length of the epigenetic genes in the genome (Supplementary Figure 2b).

Notably, we found that all the variants near the *POLR1C* gene were significantly more frequent in UKB-AFR than UKB-EUR, including rs116775335 (allele frequency [AF]; AFR = 0.230, EUR = 2.7 x10^-05^, p = 3.7 x 10^-60^) and rs7743244 (AF; AFR = 0.551, EUR = 0.061, p = 9.4 x10^-322^) (Supplementary Data 1).

The frequency disparities observed in epigenetic gene variants among UKB-AFR and UKB-EUR populations reflect a broader pattern in the genome, with frequency variations observed consistently observed across the entirety of the gene spectrum when comparing UKB-AFR and UKB-EUR populations (see Supplementary Note 2).

### AFR have significantly more common variants compared to EUR ancestry

We found that UKB-AFRs have more common epigenetic gene variants (>1% minor allele frequency) compared to UKB-EURs (Figure 2a). Many variants exhibit frequencies that are orders of magnitude higher in the UKB-AFR population than the UKB-EUR population, some by a factor of a million or more. Conversely, most variants that exhibit frequencies higher in the UKB-EUR population than the UKB-AFR population are only a few hundred to a few thousand orders (as above) of magnitude more prevalent in the UKB-EUR population (Figure 2b).

**Figure 2:**
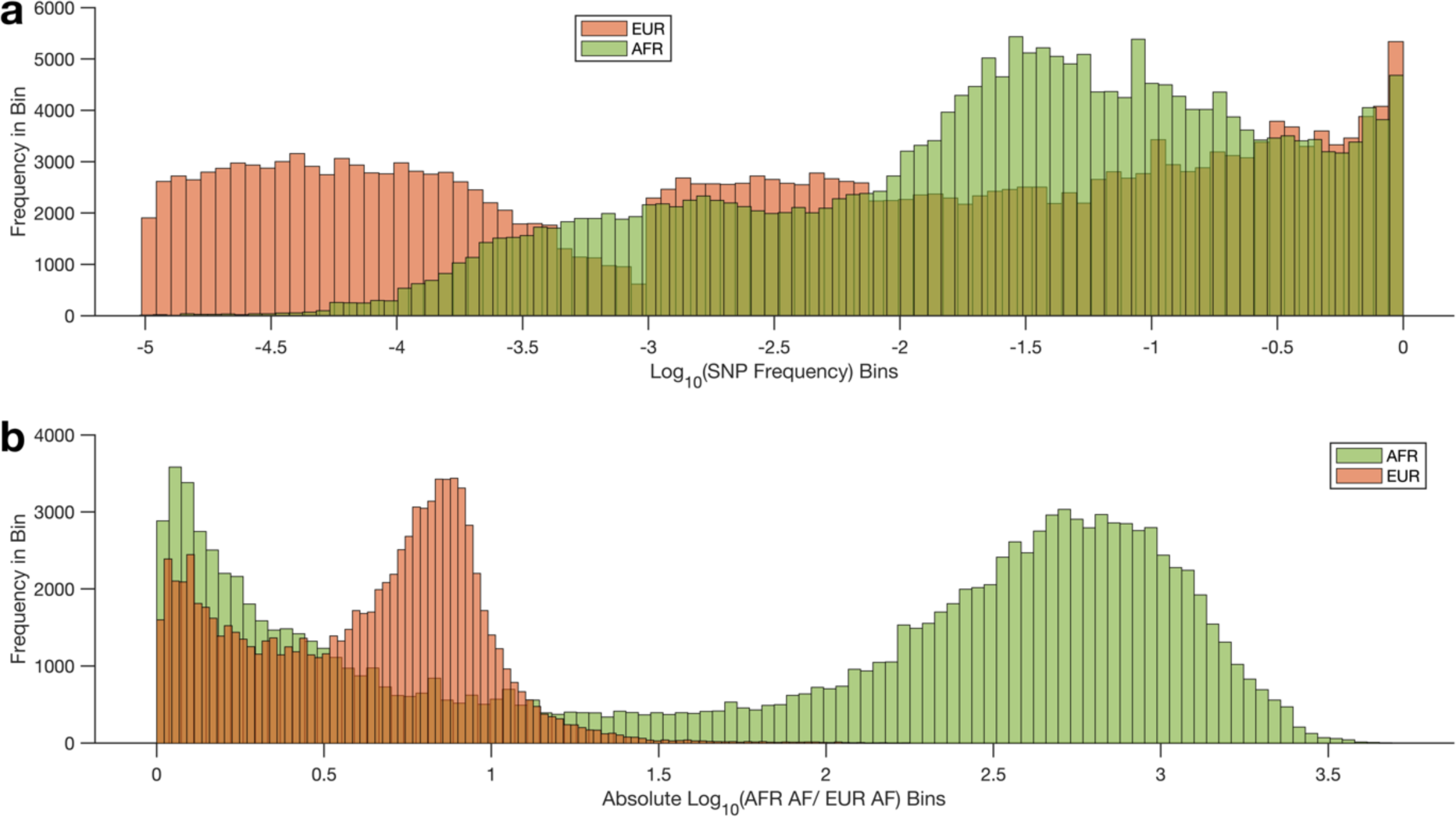
Comparative Distribution of Epigenetic Gene Variants in AFR and EUR UKB Cohorts. **(a)** Superimposed histograms illustrate the distribution of SNP frequencies for epigenetic gene variants in AFR and EUR cohorts, sorted into bins by the base-10 logarithm of their frequencies. **(b)** The histogram demonstrates the distribution of allele frequency ratios (AFR/EUR), with bins representing the absolute value of the base-10 logarithm of the frequency ratio. For detailed binning and calculation methodologies, refer to the Methods section.

The common variants that exhibited the most significant difference in frequency between UKB-AFRs and UKB-EURs were: rs4983416 in the *MTA1* gene (AFR = 0.228, EUR = 0.983, p = 9.49 x 10^-322^), rs1267502 in the *JARID2* gene (AFR = 0.252, EUR = 0.985, p = 9.63 x 10^-322^), and rs10964101 in *SMARCA2* (AFR = 0.819, EUR = 0.094, p = 9.49 x 10^-322^).

### Analysis of rare variants and their functional impact across AFR and EUR populations

Many rare variants also exhibited significant differences in frequency, including the *SP140* gene variant rs73104290, which is 4824 times more frequent in UKB-AFR (0.07) compared to UKB-EUR (1.46 x 10^-5^, p = 8.6 x 10^-188^) and a *SETD7* gene variant rs578143624, that is 178 times more frequent in UKB-EUR (0.002) than UKB-AFR (1.1 x 10^-5^, p = 2.4 x 10^-17^).

From the 123,168 variants significantly more frequent in UKB-AFR, our further analysis revealed 9,007 of these to be common in the UKB-AFR population (AF > 0.1) but rare in the UKB-EUR population (AF < 0.001), see Figure 2b. Furthermore, we conducted analyses to assess the functional impact of these 9,007 genetic variants and observed several patterns. We found that the top three sequence ontology (SO) terms for variant consequence classification by Ensembl Variant Effect Predictor (VEP) were intron variants (5,598), intergenic variants (2,019), and upstream gene variants (375), see Supplementary Figure 3a.

Furthermore, the most frequent variants were those within genes annotated for protein-coding (4,952 counts), followed by unspecified categories (N/A) with 2,287 counts (intergenic variants, whose impact on protein function and gene expression could not be predicted), and long non-coding RNA (lncRNA) with 769 counts (refer to Supplementary Figure 3b). These intron variants, although non-coding, can affect gene regulation and expression, as detailed in Supplementary Data 1.

Additionally, we employed the Ensembl Variant Effect Predictor (VEP) ^32^ to assess the functional impact of variants with reference allele frequencies exceeding 0.1 in African populations. The results revealed that a majority of these variants (6,690) were classified as “MODIFIER”, indicating that they are usually non-coding variants or variants affecting non-coding genes where predictions are difficult, or there is no evidence of impact (Supplementary Figure 3c). Interestingly, the top three most severe consequences—specifically affecting gene regulation and expression—were intron variants (5,598 instances), exemplified by rs113753107, rs17052326, and rs4234635 (see Supplementary Data 1). Notably, many variants are found within introns—non-coding segments within genes annotated for protein synthesis—which hints at their potential involvement in regulatory processes such as alternative splicing.

Overall, these results show that the frequency of SNPs located within epigenetic genes varied between UKB-AFRs and UKB-EURs, and these SNPs are more frequent in the HMT class of epigenetic genes, particularly *PRMT6*. Our findings necessitate further investigation into the biological consequences of these variants in relation to population-specific susceptibility or resistance to diseases. They underscore the importance of population-based studies in genomic medicine.

### Genetic variants discriminate between AFRs and EURs

All our previous comparisons were based on the UKB datasets of AFR and EUR. We also wanted to determine how data from AFR living in Africa compared to that from the UKB-AFR population so that we could relate further analyses based on the UKB-AFR population to AFR in Africa. We obtained whole genome sequence datasets from the H3Africa Project ^33,34^, Human Genome Diversity Project ^35^, and the 1000 Genomes Project ^14^, for details on the specific populations and the sample sizes, see Supplementary Data 1. Our analysis was explicitly tailored to investigate variants that were common across all sampled populations, focusing on shared genetic variation as detailed in the Methods section. Therefore, we excluded very rare variants (AF < 1 x 10^-5^) unique to individual groups to avoid potential biases in detecting low-frequency variants and to maintain statistical robustness. This selective approach was crucial in ensuring consistency and reproducibility of our findings across diverse populations. Integrating the UKB SNP frequencies with those from these projects and employing unsupervised hierarchical clustering, we observed expected segregation patterns: African populations clustered apart from Europeans based on the reference allele frequency of variants in epigenetic genes, with those in the UKB aligning closely with the respective continental datasets (Figure 3a and Supplementary Figure 4).

**Figure 3:**
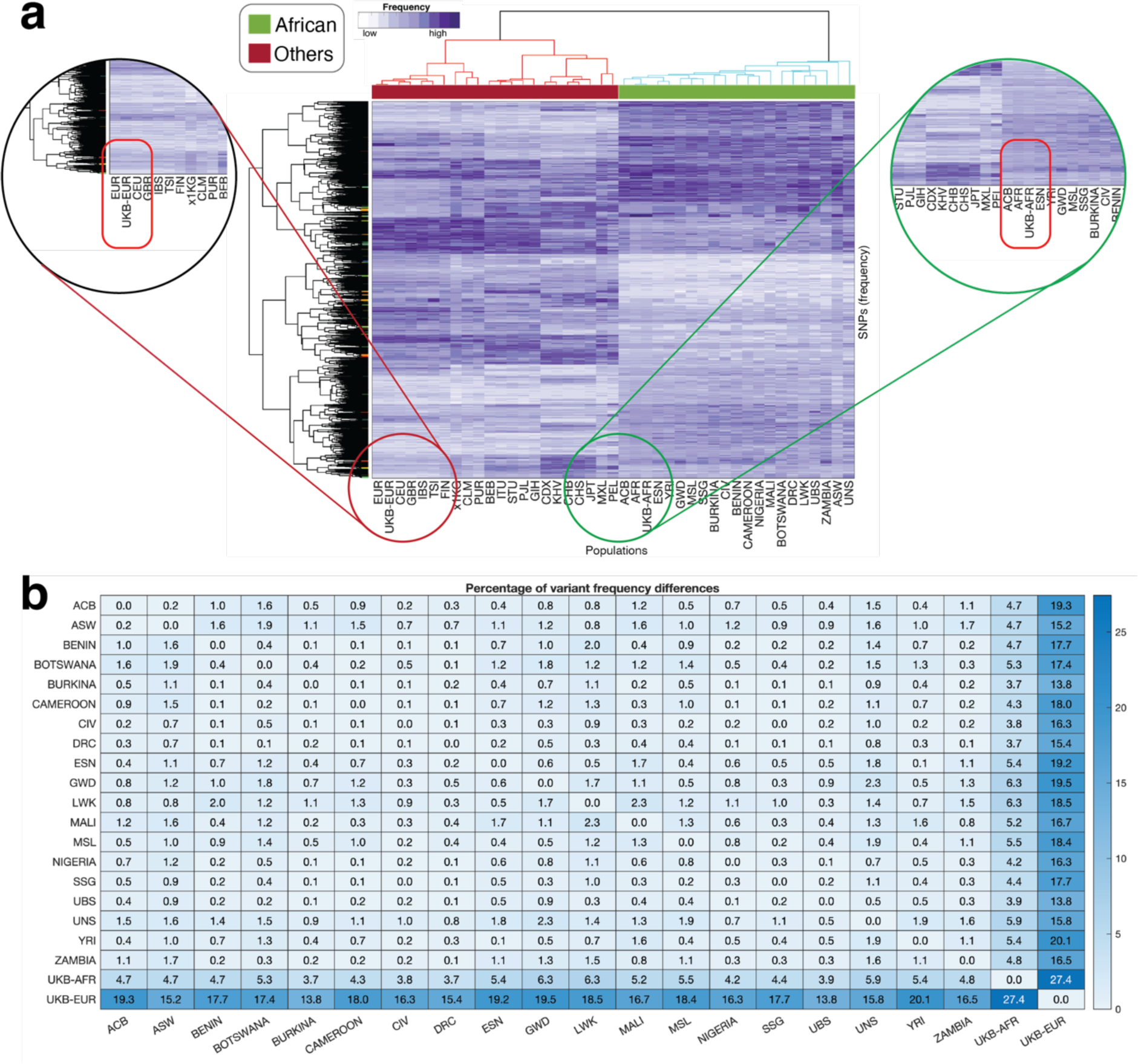
**(a)** Clustering of various populations based on the allele frequencies of SNPs at epigenetic gene loci. Increasing colour intensities denote higher frequencies of SNPs in the populations. The clustergram was produced using unsupervised hierarchical clustering with the Euclidean distance metric and complete linkage. **(b)** Percentage of variants that demonstrate a statistically significant difference between each population as determined using the Fisher exact test. The UKB-AFR and UKB-EUR represent the UK Biobank populations. See Supplementary Data 1 for the number of samples genotyped for each population.

To further delve into the genetic distinctions between AFRs residing in Africa and individuals of AFR ancestry outside of Africa and their divergence from the EUR population in the UKB, we systematically compared the frequencies of SNP variants across these groups. Employing Fisher’s Exact Test, we scrutinised each SNP for significant frequency differences between each pair of subpopulations.

The median percentage of SNPs exhibiting significant frequency variation amongst all the AFR subpopulations in the study was 0.53% (range: 0.04% - 2.33%), as depicted in Figure 3b (also see Supplementary Figure 5). In contrast, the median percentage of SNPs differing significantly in frequency between the AFR populations from Africa and the EUR population in the UKB was markedly higher, at 17.4% (range: 13.8% - 20.1%), similar to the comparison between UKB-AFR and UKB-EUR population.

Unsurprisingly, our analysis revealed a compelling genetic similarity between AFRs living in Africa and those of UKB-AFR ancestry, with their SNP frequencies demonstrating a strong correlation. The proportion of variants with significantly different frequencies ranged from 3.7% between the UKB-AFR population and the Burkina Faso population to 6.3% between the UKB-AFR population and the Luhya people in Webuye, Kenya. Moreover, this subset of the AFR population bore genetic variants congruent with those observed in broader AFR populations. This similarity is expected, as many AFR individuals in the UKB are either recent immigrants or have ancestry only a few generations back from Africa.

The SNP frequencies observed in the EUR subset of the UKB mirrored patterns previously documented in genome-wide analyses of EUR populations. Crucially, the similarity between the UKB-AFR cohort and the AFR population from the 1000 Genomes Project (shown as in clustergram AFR-AF) and the congruence between AFR subpopulations, as depicted in Figure 3 and Supplementary Figure 4, provided the impetus to utilise the UKB-AFR population as a representative dataset for discerning epigenetic gene variant differences between EUR and AFR populations. Additionally, the UKB datasets include phenotype data that was necessary for a trait analysis.

### Traits and diseases associated with variants at epigenetic gene loci

To explore the relevant traits associated with the variants located near or in the genome sequences that encode epigenetic genes, we searched the GWAS Catalog (https://www.ebi.ac.uk/gwas) for previously reported phenotypes associated with epigenetic variants, extended to include variants in moderate LD (r2 ≥ 0.50) with the GWAS hits (see the Methods sections). We discovered that 755 epigenetic variants were associated with 373 traits. The most prominent traits included sex hormone-binding globulin level (SHBG), waist circumference, and height, which were associated with 32, 24, and 8 variants (Figure 4a), respectively. Notably, of the 32 variants linked to SHBG, 8 were significantly more prevalent in UKB-AFR and 24 in UKB-EUR, while, among the 24 variants associated with waist circumference, 4 were found more frequently in UKB-AFR and 20 in UKB-EUR. For a complete list of traits, genes, and genetic variants reported in the GWAS Catalog that vary between UKB-AFR and UKB-EUR, see Supplementary Data 2.

**Figure 4:**
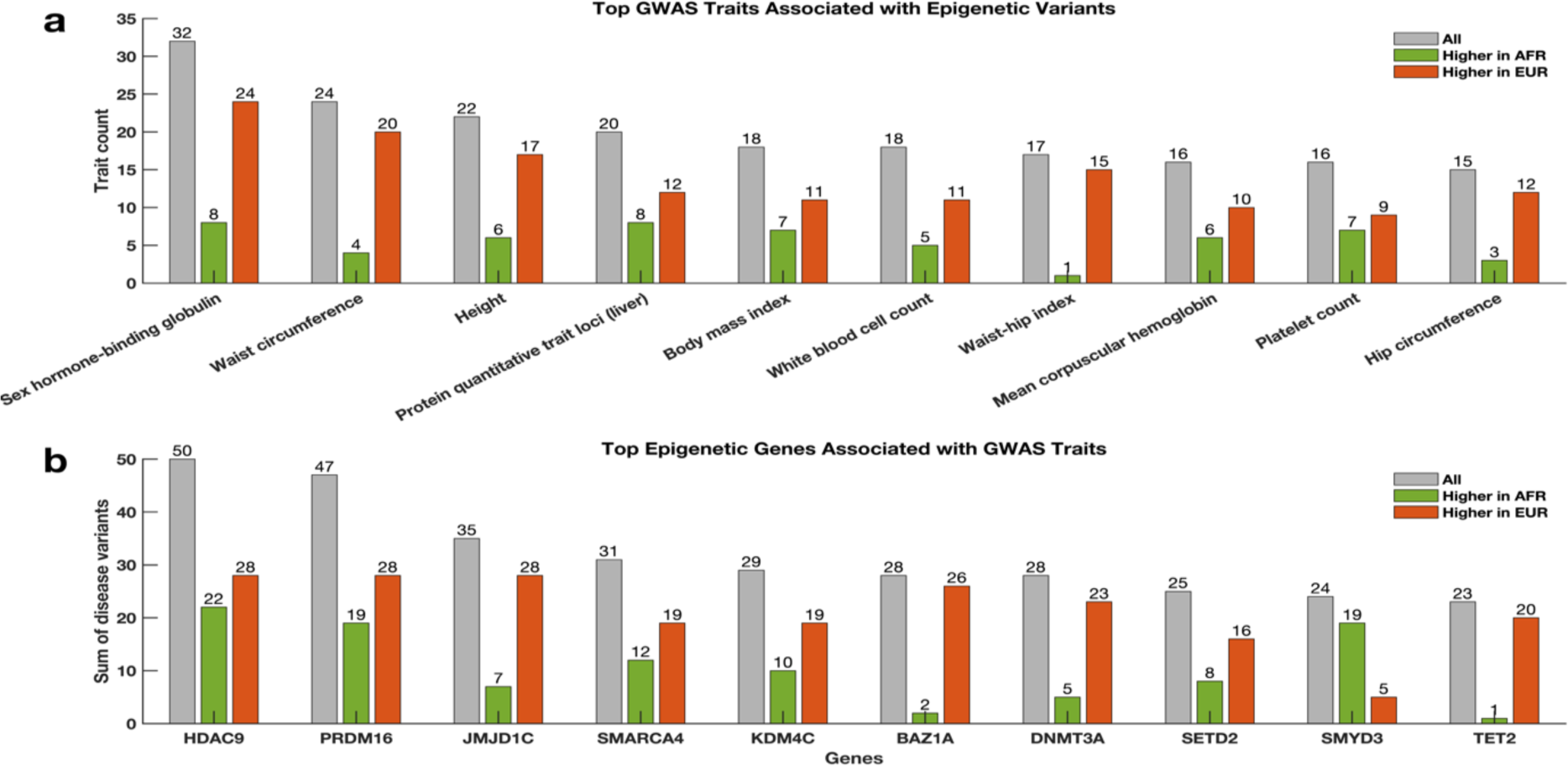
Distribution of GWAS Catalog terms for epigenetic gene variants. (a) Bar graph of the top-10 ranking traits associated with epigenetic gene variants. (b) Bar graph of the top-10 ranking epigenetic genes most associated with traits in the GWAS Catalog. The bar graphs are coloured based on the significance of variants in AFR and EUR. The orange bar depicts variants significantly more frequent in EUR, green for variants significantly more frequent in AFR, and grey for variants not significantly different between AFR and EUR populations.

We also analysed the epigenetic genes most correlated with these traits reported in the GWAS Catalog. The gene *HDAC9* was associated with the greatest number of traits, totalling 50 variants, followed by *PRDM16* with 47 variants and *JMJD1C* with 35 (Figure 4b). Of the variants located in or near the *HDAC9* gene, 22 were significantly more frequent in UKB-AFR and 28 in UKB-EUR. Similarly, for *PRDM16*, 28 variants were significantly more frequent in UKB-EUR and 19 in UKB-AFR.

Here, we show that most of the epigenetic gene variants associated with various traits in the GWAS Catalog differ in frequency in AFR and EUR. By extension, these differences may impact the extent to which a particular locus is found associated with a trait and the occurrence of the disease or trait in each population.

### Quantitative trait impact of variants at epigenetic gene loci

We evaluated the functional impact of epigenetic gene variants that cause changes in DNA methylation (methylation Quantitative Trait Loci [mQTLs]), gene expression (expression Quantitative Trait Loci [eQTLs]), transcript splice site (splice Quantitative Trait Loci [sQTLs]), and histone post-translational modifications (enhancer histone Quantitative Trait Loci [hQTLs]). Here, 908 variants among the 223,766 variants (0.41%) were eQTLs in various tissues (Supplementary Figure 6a). We cross-referenced the eQTL variants in the epigenetic genes to the GWAS Catalog and showed that, among the 908 eQTLs, 24 were reported in the GWAS Catalog as being associated with various traits (Figure 5a). For example, the *GSK3B* eQTL variant rs189174, associated with serum alkaline phosphatase levels^36,37^ is 2.7 times more frequent in UKB-AFR (0.91) compared to UKB-EUR (0.34). Also, the *KDM2A* eQTL variant rs12785905, which is 10 times more frequent in UKB-EUR (0.06) compared to UKB-AFR (0.006), is associated with prostate cancers ^38,39^.

**Figure 5:**
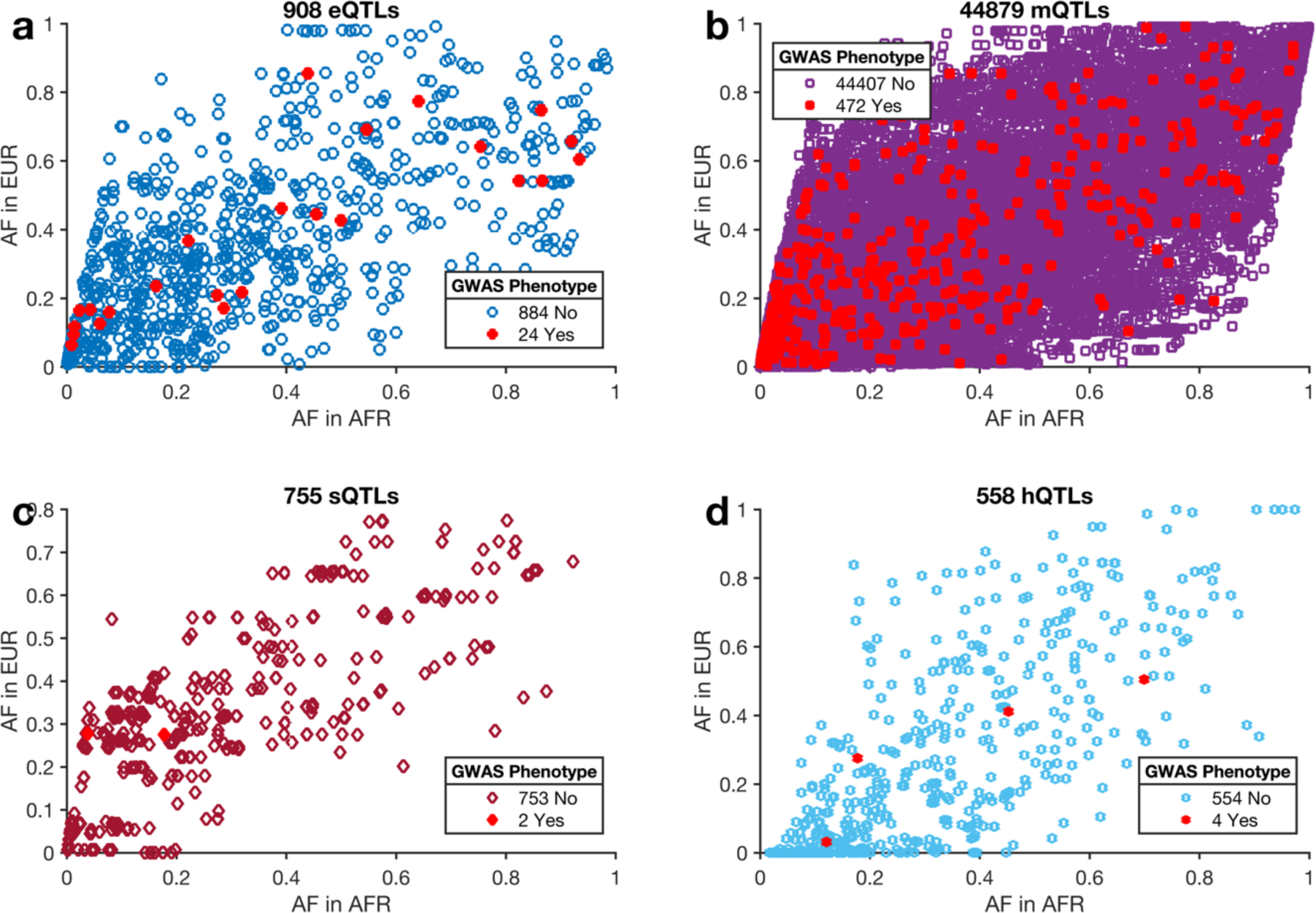
The frequency of quantitative trait loci of epigenetic genes in AFR versus EUR in the UKB. Frequency of (a) eQTL, (b) mQTL, (c) sQTL, and (d) hQTL variants among AFR and EUR. The red-coloured points show QTL variants associated with traits or diseases in the GWAS Catalog. An interactive visualisation of Figure 5 can be found <u>here</u>.

In addition, we found 44,879 mQTLs out of the 223,766 (20.1%) variants at epigenetic gene loci (Supplementary Figure 6b), of which, 472 are reported to be associated with various traits (Figure 5b). The frequency of 96% of the mQTLs differs significantly between UKB-AFR and UKB-EUR, a finding that may influence the penetrance of traits associated with the two populations ^40^ (Figure 5b and Supplementary Figure 6b). For example, the *SETD8* gene mQTL variant rs28577594 is associated with high-density lipoprotein cholesterol levels ^41,42^ and is 4.2 times more frequent in UKB-EUR (0.723) than UKB-AFR (0.171). Conversely, the *BRD1* gene variant rs763236 is associated with DNA methylation variation ^43^ and is 33.7 times more frequent in UKB-AFR (0.363) compared to UKB-EUR (0.011).

Furthermore, we found 775 (0.35%) sQTLs among the 223,766 variants (Supplementary Figure 6c), of which 2 variants are associated with various traits in the GWAS Catalog (Figure 5c). These sQTLs include rs12575252 (AF; AFR = 0.54, EUR = 0.35, p = 1.4 x 10^-194^) of the *TRIM66* gene associated with waist-hip ^44^ and rs4833687 of the *PRDM5* gene, associated with type 2 diabetes mellitus ^45^, which is 7 times more frequent in UKB-EUR (AFR = 0.04, EUR = 0.28, p = 9.5 x 10^-322^).

Finally, we found 558 (0.17%) hQTLs (Supplementary Figure 6d), among which 4 are associated with traits (Figure 5d). For example, the *GSK3B* gene hQTL variant rs189174 is associated with alkaline phosphatase levels and is 2.7 times more frequent in UKB-AFR (0.91) compared to UKB-EUR (0.34), and the *NSD1* gene hQTL and mQTL variant rs7724386 that is associated with the neutrophil count is 2.2 times more frequent in UKB-EUR (0.58) compared to UKB-AFR (0.26).

We hypothesize that when QTLs with different frequencies across populations (e.g., AFR and EUR in this case) are linked to traits or diseases, they will likely influence the prevalence or penetrance of those traits in different populations ^46^ (Supplementary Figure 6 and Supplementary Figure 7).

### Variants exhibit multiple quantitative trait loci effects and disease associations

We found that 632 (70.5%) of the identified eQTLs also function as mQTLs for the same gene, 43 (4.8%) overlap with sQTLs, and 4 (0.4%) coincide with hQTLs (Supplementary Figure 8 and Supplementary Figure 9). Furthermore, we found a significant overlap among the various QTLs, which may indicate an overlap of variant effects on expression, DNA methylation, mRNA splicing, and histone mark modification. Our findings suggest a substantial degree of multi-scale regulation of gene expression, methylation, and histone accessibility at the transcription level. For instance, variants that change methylation, chromatin accessibility, or transcription factor binding at an enhancer location for a specific gene can result in overlapping mQTLs, hQTLs, and eQTLs, impacting transcription, promoter/chromatin accessibility, and gene accessibility ^47–50^.

To explore the relevance of regulatory variation affecting epigenetic gene QTLs in traits and disease, we assessed the overlap between QTLs and the GWAS Catalog, extended to include variants in high linkage disequilibrium (r² ≥ 0.50) with GWAS hits. Our findings demonstrate a significant enrichment of QTLs (1.1% of associations in the GWAS Catalog) compared to non-QTLs (0.16% of associations), highlighting their association with a broad array of GWAS traits and diseases (odds ratio [OR] = 10, p = 3.0 x 10^-144^). Among the different classes of QTLs, mQTLs showed the most substantial association odds (OR = 6.8, p = 6.9 x 10^-144^), followed by eQTLs (OR = 17.4, p = 3.4 x 10^-21^), hQTLs (OR = 4.6, p = 0.012), and sQTLs (OR = 1.7, p = 0.33), as presented in the updated Table 1. These results underscore the significant impact of eQTLs and mQTLs in modulating trait associations, confirming their predominant role among QTL classes.

**Table 1:**
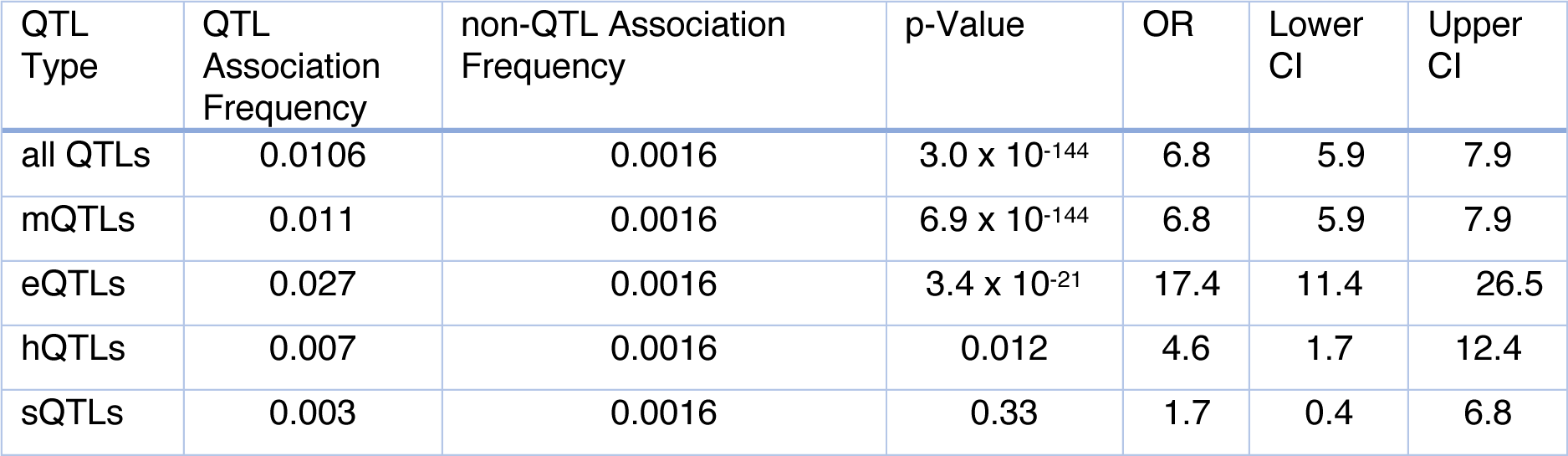
Comparative Associations Between QTLs and GWAS Traits.

### Variant frequencies and QTLs to determine the potential for biomarker discoveries among AFR and EUR

To explore the relevance of epigenetic gene variants that are eQTLs, we assessed the association of the variants with 28 blood and urine biomarker tests in the UKB separately for AFR and EUR (see Methods section). These biomarkers, including albumin, aspartate aminotransferase, cholesterol, and glycated haemoglobin, were selected based on their established significance in reflecting metabolic, liver, and kidney functions. This choice allows for a nuanced understanding of the potential impact of epigenomic variants on a broad spectrum of physiological processes, facilitating a more detailed exploration of their implications in disease susceptibility and population-specific health outcomes (see Methods section, Supplementary Figure 9).

Overall, our analysis revealed 26,564 variants significantly associated with biomarker traits across both populations (Supplementary Figure 10 and Supplementary Data 3). Unsurprisingly, we showed a bias in the results for each population towards variants that are most frequent in that population (Figures 6a and 6b). Specifically, all 17 variants (100%) that were uniquely and significantly associated with traits in the UKB-AFR population were also significantly more frequent in UKB-AFR. In contrast, among the 16,722 variants (approximately 62%) were significantly more frequent in UKB-EUR individuals, while 8,079 variants (approximately 33%) were significantly more frequent in UKB-AFR (Figure 7c). Among the 290 variants found to be significant in both groups, 214 (approximately 75%) were significantly more prevalent in UKB-EUR, and 73 (approximately 25%) were more frequent in UKB-AFR, with only 3 showing no particular frequency bias (Figure 6d). This detailed distribution underscores significant genetic differences between populations, potentially impacting the biomedical relevance of these variants.

**Figure 6:**
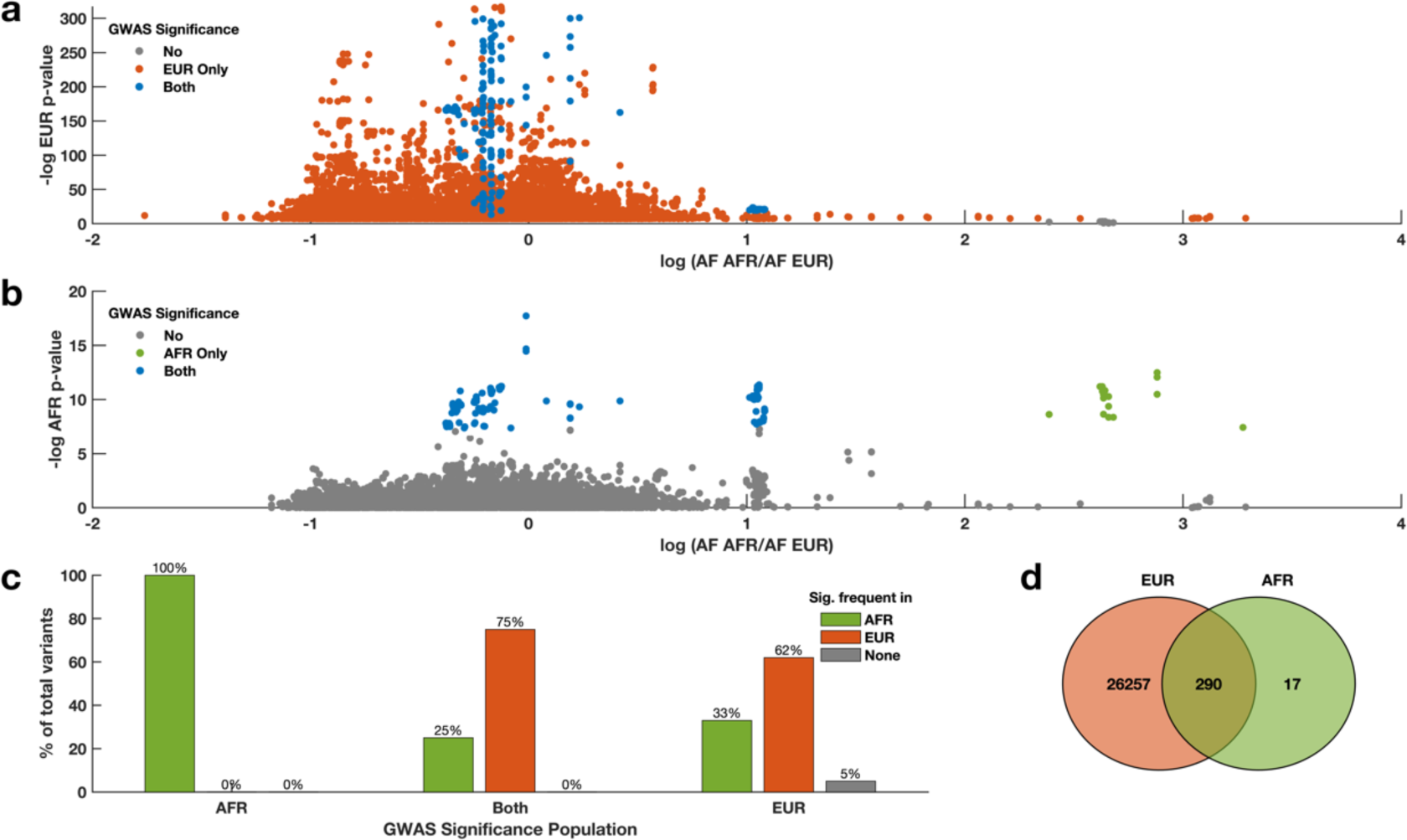
Significant association of SNPs with biomarkers in relation to the frequency of SNPs in AFR and EUR. (a) Frequency of variants in AFR/EUR vs. the −log10 p-value in EUR. This scatter plot shows the distribution of SNPs based on their allele frequency ratios (AFR/EUR) on the x-axis and their statistical significance in EUR populations (-log10 p-value) on the y-axis. The colours show the statistical significance: blue for variants significantly associated with biomarker levels in both AFR and EUR, orange for significant associations only in EUR, and grey for non-significant variants in EUR. (b) Same as (a) but showing the −log10 p-value in AFR. Colours indicate statistical significance: blue for variants significantly associated with biomarker levels in both AFR and EUR, green for significant associations only in AFR, and grey for non-significant variants in AFR. (c) Percentage of variants significantly associated with biomarker traits, grouped by their significance in AFR, EUR, or both groups. The colours show details about statistical significance: orange for variants significantly more frequent in EUR, green for variants significantly more frequent in AFR, and grey for variants with similar frequencies in AFR and EUR. This panel helps illustrate the distribution of significant variants across populations. (d) Venn diagram showing the overlap of significant variants between EUR and AFR populations. The red circle represents variants significant in EUR, the green circle represents variants significant in AFR, and the overlap shows variants significant in both populations.

**Figure 7:**
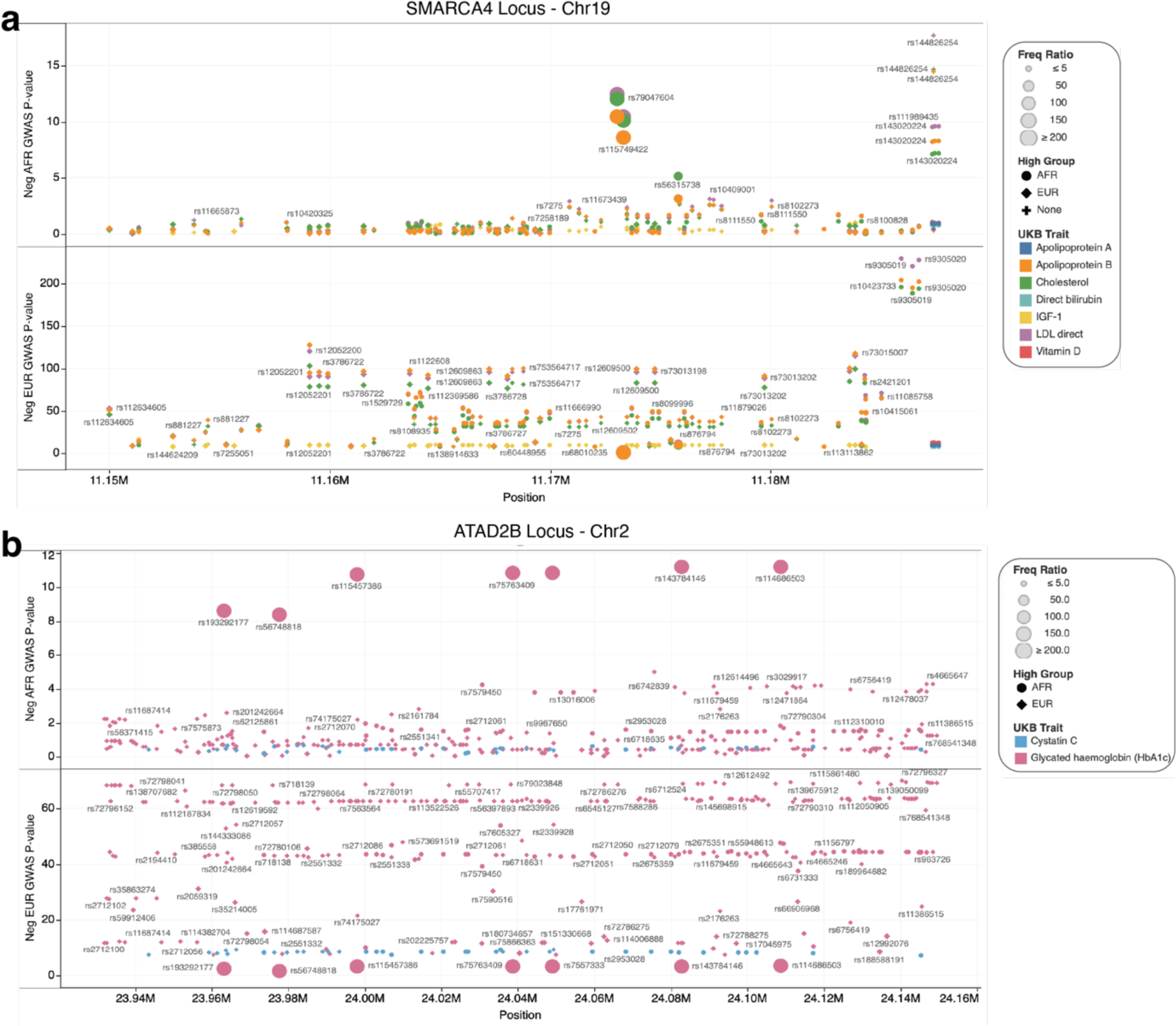
Chromosomal regional plots at the SMARCA4 and ATAD2B gene loci. (a) SMARCA4 chromosomal positions vs base-10 negative logarithm GWAS-p-values for biomarker traits associated with AFR and EUR. The chromosomal positions are filtered, ranging from 11,150KB to 26,845KB of chromosome 19. (b) ATAD2B chromosomal positions vs base-10 negative logarithm GWAS-p-values for biomarker traits associated with AFR and EUR. The chromosomal position filter ranges from 23,000KB to 24,300KB. The colours show details about UKB biomarker traits, whereas the marker sizes show base 10 absolute logarithms of SNP frequency in AFR vs EUR. The shapes show details about the group in which variants are significantly more frequent. The marks are labelled by the variants’ SNPdb IDs. Interactive visualisation of Figures 7a and 7b can be found <u>here</u>.

Insulin-like growth factor-1 (*IGF-1*) was identified with 2,864 variants, showing a significant association prevalence of 10.77% among the biomarker traits studied. Of these, 2,860 variants were found only in the UKB-EUR population, 4 were shared between both populations, and none were exclusive to the UKB-AFR population.

This was followed by Sex Hormone Binding Globulin (SHBG) with 1,960 variants (7.39%), of which 1,550 were more frequent in UKB-EUR, 408 were shared between both populations, and 2 were found only in UKB-AFR. Alkaline phosphatase also showed a high number of associations with 2,017 variants (7.58%), where 1,984 variants were more common in UKB-EUR, 33 shared, and none exclusive to UKB-AFR (see Supplementary Figure 9 and Supplementary Figures 11a to 11c).

Among some of the notable discoveries was rs1122608, a *SMARCA4* gene mQTL variant, which we found to be associated with cholesterol (GWAS p = 4.1 x 10^-82^), apolipoprotein B (p = 3.2 x 10^-98^), low-density lipoprotein (LDL; p = 1.2 x 10^-94^), and IGF-1 (p = 1.2 x 10^-9^) in UKB-EUR only, and not replicated in UKB-AFR. Interestingly, rs1122608 was 7.7 times (Fisher’s p-value = 9.5 x 10^-322^) more frequent in UKB-EUR (AF = 0.254) than in UKB-AFR (AF = 0.033), as illustrated in Figure 7a and Supplementary Figure 12 and Supplementary Figure 13.

Additionally, at the loci in the ATAD2B gene on chromosome 2, we identified several variants associated with Glycated haemoglobin (HbA1c) only in UKB-AFR. The lead variant at this locus, rs114686503 (GWAS p-value = 6.43 x 10^-12^), was 425.91 times more frequent in UKB-AFR compared to UKB-EUR, providing a compelling insight into the population-specific prevalence of these SNPs associated with the traits (Figure 7b). Furthermore, nearby variants in strong LD with rs114686503, including rs75763409 (GWAS p-value = 1.4895 x 10^-11^, 438.5 times), rs7557333 (GWAS p-value = 1.4747 x 10^-11^, 438.39 times), and rs115457386 (GWAS p-value = 1.7479 x 10^-11^, 426.57 times), were also significantly more frequent in UKB-AFR than UKB-EUR, yet they were not associated (directly or by replication) with any traits in UKB-EUR (see Supplementary Figure 14 and Supplementary Data 3).

We found several QTLs associated with various traits whose frequencies differ between the UKB-AFR and the UKB-EUR, and within the AFR populations (Figure 8a). For example, at the *KDM2A* gene loci on chromosome 11, the eQTL and mQTL variant rs12785905 is 10 times more frequent in the UKB-EUR than in the UKB-AFR and was associated with IGF-1 levels in the UKB-EUR (GWA p-value = 4.0 x 10^-16^), but not replicated in the UKB-AFR (replication p-value = 0.09). Near the *KDM2A* locus, all other variants in strong LD (R^2^ > 0.6) with rs12785905, including rs35826789 (8 times), rs56088284 (9 times), rs34560402 (10 times), and rs12785906 (10 times), are significantly more frequent in the UKB-EUR and associated with IGF-1 levels in the UKB-EUR only. As shown in Figure 8a, there is also significant variation between different AFR populations. For example, the mQTL rs2236682 (associated with creatinine levels in the UKB-AFR and the UKB-EUR) ranges in frequency from approximately 0.15 in the Yoruba population (YRI) to 0.49 in the Zambian population. The frequency in Asian populations is 0.2, and EURs is 0.37 according to dbSNP, which has the AFR frequency as 0.29. For clinical use, the intercontinental variation may be important to consider.

**Figure 8:**
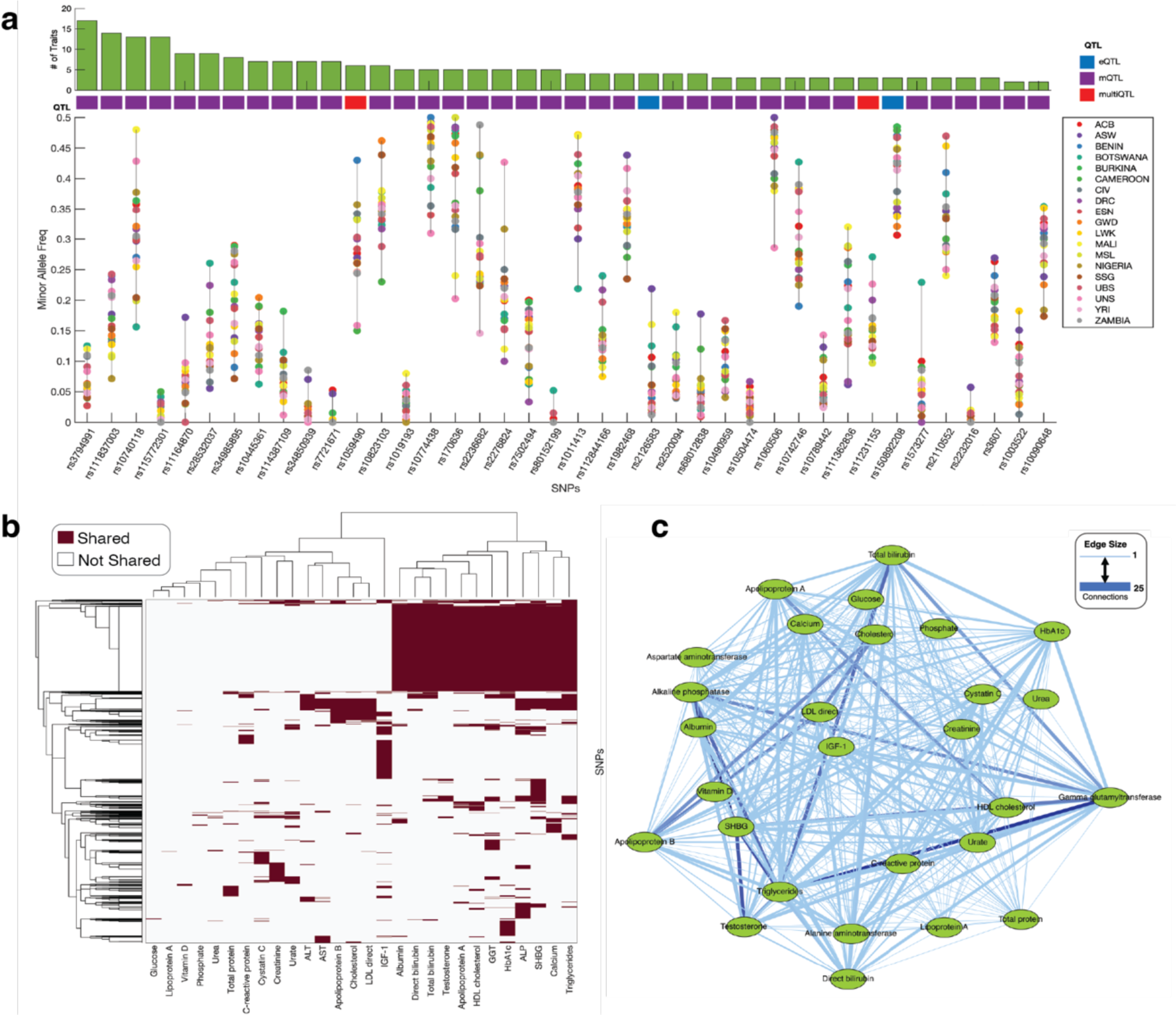
Within African populations, variations in the frequency of minor alleles for lead QTL SNPs and the genetic correlation of biomarker traits are evident. Specifically, within a 2 MB region encompassing QTLs linked to key biomarker traits, the minor allele frequencies across various African populations highlight the continent’s extensive genetic diversity. Each point corresponds to the frequency of a minor allele in a particular population for a given SNP. Details of the AFR populations can be found in Supplementary Data 1. (b) Unsupervised hierarchical clustergram of blood and urine biomarkers in the UKB based on the GWA calculated p-value of associations. The darker colours show statistically significant associations with the variation along the rows for the biomarkers coloured along the columns. (c) Connectivity network of biomarker traits based on shared GWA variants.

Furthermore, we found that clusters of loci associated with various biomarker traits in the UK Biobank are significantly enriched for QTLs. Among the 26,564 associations, we identified 19,102 (71.9%) as mQTLs, 898 (3.38%) as multi-QTLs, 63 (0.24%) as eQTLs, 30 (0.11%) as sQTLs, 8 (0.03%) as hQTLs, and 6,463 (24.33%) did not fall into any QTL category (Supplementary Figure 15). For example, at the loci in and near the *SMARCA4* gene, among the unique 300 significant variants associated with 7 traits, 211 are mQTLs, and 13 are multi-QTLs (Supplementary Figure 16a). Furthermore, at the loci in or near the ATAD2B gene, among the 356 significant variants associated with 2 traits (Supplementary Figure 16b), we found 270 mQTLs and 6 multi-QTLs. These associations included 270 unique mQTLs, 6 unique multi-QTLs, and 41 non-QTLs.

Our analysis reveals that loci associated with biomarker traits demonstrate a significant enrichment for various types of QTLs. Specifically, among the 26,564 evaluated associations, we identified a predominant presence of mQTLs (19,102 or 71.9), suggesting a potential link to DNA methylation dynamics. However, it is important to note that our conclusions are derived from allele frequency differences and the association of these variants with mQTLs rather than direct observation of DNA methylation changes. This indirect evidence suggests that the genetic variants we identified may influence gene regulation, possibly through mechanisms including but not limited to DNA methylation and/or other epigenetic processes (Supplementary Figures 14 and 15a-b).

### Biomarker traits associated with epigenetic variants are strongly correlated

Next, we evaluated the correlation between traits associated with epigenetic gene variants. Here, we found a cluster of loci shared across traits, including blood calcium levels, total bilirubin, and albumin (Figure 8b). For example, we found that the *GTF2H4* gene mQTL variant rs1264313 is associated with the levels of 17 biomarkers, including albumin, calcium, and cholesterol (Supplementary Figure 16). Furthermore, the *ARID1A* gene variant rs114165349 is associated with variations in the serum levels of 17 biomarkers. See Supplementary Figure 17 for a list of some of the variants associated with the serum levels of multiple biomarkers. We find that the biomarkers with the greatest number of shared associations are alkaline phosphatase and SHBG (864 shared variants), gamma-glutamyl transferase and triglycerides (808 variants), and SHBG and triglycerides (718 variants), see Figure 8c and Supplementary Data 4.

Interestingly, we observed that certain traits do not share a significant number of epigenetic variants. For instance, variants associated with glucose levels are not commonly shared with those linked to HbA1c. Similarly, variants influencing urea levels do not overlap extensively with those affecting albumin and bilirubin levels. This lack of shared variants suggests that these traits are influenced by distinct genetic and epigenetic mechanisms, highlighting the complexity of gene-trait associations.

## Discussion

We analysed the frequencies, associated traits, and the functional impact of epigenetic gene variants among UKB-AFR and UKB-EUR ancestry individuals. We found that 88.4% of SNPs at epigenetic gene loci differed in variant allele frequencies between UKB-EUR and UKB-AFR. Previous studies have reported that the genetic variants associated with various phenotypes differ among individuals of different ancestry ^41,42^. We found *PRMT6,* an HMT class epigenetic gene, to have the largest number of variants, many of which differed significantly in frequency between UKB-AFR and UKB-EUR. *PRMT6* encodes arginine methyltransferase 6, which is involved in a variety of biological processes such as cell death, cell cycle progression, and RNA processing ^51,52^, and the gene has been linked with many traits and diseases, including serum protein levels ^53^, and rapid progression of HIV/AIDS ^54^.

We showed that many common (AF > 0.1) epigenetic gene variants in UKB-AFR are rare in UKB-EUR individuals. There is increasing evidence that the allelic spectrum of risk variants at a given locus might include rare, low-frequency ^55^, and common genetic variants ^56^. Such a scenario may imply that traits associated with these common variants are biased towards the population with high-frequency variants, as the shared variant predisposes to a common disorder or trait ^57^. While AFR populations display a rich mosaic of genetic diversity, characterised by extensive substructure and reduced linkage disequilibrium compared to EUR populations, our findings highlight that, despite these within-group variances, the differences in the frequencies of epigenetic gene variants between AFR sub-populations are less marked than those observed between AFR and EUR groups as a whole ^58^. This observation suggests a foundational level of genetic similarity among various AFR sub-populations, which contrasts with the more pronounced differences observed when comparing AFR populations with EUR ones, reflecting both the deep genetic diversity within AFR and the distinct evolutionary paths that have shaped these populations.

Our analysis of prior GWAS findings reveals that variants in epigenetic genes are predominantly linked to physical attributes such as height, weight, and BMI, as well as haematological metrics like white and red blood cell counts and haemoglobin levels. The above-listed traits have previously been linked to epigenetic regulation and change ^59,60^. Additionally, nearly 70% of these variants we identified as associated with traits in the GWAS Catalog and UKB biomarker measures are enriched for QTLs. Whereas previous studies showed that disease variants are enriched for eQTLs, we show that most eQTLs are mQTLs ^49,50^ and that disease traits are enriched for mQTLs, i.e., the significant functional impact mechanism associated with traits could be variations in DNA methylation due to SNPs. Our assumption is supported by recent findings that genetic variants that modify DNA methylation are a dominant mechanism through which genetic variation leads to gene expression differences among humans since a substantial fraction (16%) of mQTLs and sQTLs are also associated with variation in the expression levels of nearby genes (that is, these loci are also classified as eQTLs) ^48,61^. Our results show that 70.5% (632 out of 896) of eQTL variants in epigenetic genes can be explained by changes in DNA methylation, i.e., mQTLs (see Supplementary Figure 6), demonstrating the pervasiveness of co-regulated expression and methylation in the human genome. This suggested that variations in the frequency of SNPs associated with these traits could influence the detection of associated SNPs in different populations through GWAS ^11,15,16^.

We posit that SNPs prevalent in UKB-AFR populations yet infrequent in UKB-EUR might represent unstudied QTLs, overlooked in large-scale sequencing projects like the 1000 Genomes and GTEx due to their rarity in global datasets ^62^. The detection of QTLs, including eQTLs, is inherently influenced by SNP frequencies within the examined population, where SNPs less common in a global context may be inadequately characterised as QTLs ^63^. This phenomenon underscores the importance of exploring genetic diversity within specific populations to uncover distinct regulatory variants. For instance, the study by Esoh et al. identified population-specific eQTLs in African populations, emphasizing the need for targeted analysis of under-represented populations to fully elucidate genetic influences on traits and diseases ^62^.

Moreover, the identification of eQTLs presents additional complexities due to their tissue-specific nature. The regulatory influence of eQTLs depends on the tissue-specific expression of transcription factors and the accessibility of chromatin, factors which vary significantly across different tissues ^64,65^. This tissue specificity highlights the necessity for matched whole-genome sequencing and transcriptome datasets within AFR populations to identify eQTLs effectively. However, such matched datasets are scarce, particularly for AFR populations, posing a significant challenge in accurately mapping eQTLs and understanding their functional impact. This scarcity, combined with the tissue-specific expression patterns of eQTLs, emphasizes the critical need for more comprehensive genomic and transcriptomic analyses tailored to diverse populations.

Our findings show how certain traits may be genetically linked, sharing underlying epigenetic mechanisms that influence their expression ^66–70^. This suggests that variations in epigenetic genes can concurrently affect multiple traits, revealing a complex network of genetic interactions and epigenetic regulation contributing to trait diversity. However, in this study, we refrain from making conclusive causal inferences about these variants and focus only on the likely discovery of these variants based on the frequency of variants in each population. As a caveat, it should be noted that previous GWAS studies and the UKB have a minimal representation of AFR individuals. Hence, the distinctions delineated herein warrant cautious interpretation, acknowledging the possibility that our statistical power may be insufficient to discern many associations within African cohorts, thereby possibly omitting commonalities across the examined groups ^40,71^. Assessing our results with a bigger sample of people of AFR ancestry would be interesting. In summary, our findings show that epigenetic gene variants associated with traits tend to differ in frequencies among AFR and EUR, which impacts GWAS discoveries, and variants associated with traits are enriched for QTLs.

## Limitations

### Analysis of QTLs

While our study provides significant insights into the distribution and impact of various QTLs across the UKB-AFR and UKB-EUR populations, it is essential to acknowledge certain limitations. The QTL data utilised in our analysis were primarily sourced from databases with a significant representation of EUR ancestry individuals, primarily GTEx. Consequently, our findings’ direct applicability and validation in AFR populations remain limited. Furthermore, it is important to note that QTLs, including eQTLs, are tissue-specific, which may affect the interpretation of our results (Supplementary Figure 18). These limitations highlight the critical need for further research involving diverse populations and a range of tissue types to fully understand the complex interplay between genetics and epigenetics and their contribution to human health and disease.

## Methods

We analysed a UK Biobank (UKB) ^21^ dataset of 383,471 individuals categorised under EUR ancestry, including those identified as White, British, Irish, and individuals with "any other white background", and 5,978 individuals identified as having a recent AFR ancestry. The demographics of the UKB participants are extensively described elsewhere ^21^.

Additionally, in the UKB, ancestry categorisation commenced with participants’ self-identification. This initial classification was further refined using principal component analysis (PCA), which was succeeded by a random forest algorithm applied to the PCA results. This process enabled the reassignment of individuals to their self-identified ancestries based on a membership posterior probability greater than 0.5. Those individuals whose posterior probability did not exceed this threshold for any specified ancestry group were excluded from subsequent analyses ^21^. It is imperative to note that the categorisation of these populations in our study adheres to the UKB’s standards, which we acknowledge may not fully encapsulate the complex mosaic of genetic, cultural, and historical factors that contribute to an individual’s identity.

Our analysis focused on genotyping array data of imputed SNPs and GWAS summary statistics for various plasma biomarker tests, such as albumin, cholesterol, triglycerides, and serum enzyme levels, including alkaline phosphatase. We ensured a comprehensive evaluation of the genetic influences on these biomarkers across populations by including only high-quality variants ^21^. These variants are defined as PASS variants in gnomAD ^72^ and exhibit consistent frequencies with each population in gnomAD (AFR, AMR, EAS, and EUR frequencies are within 2-fold or have a chi-squared p-value of the difference > 1 x 10^-6^), with a frequency greater than 1 x 10^-5^ in both UKB-AFR and UKB-EUR (Figure 9).

**Figure 9:**
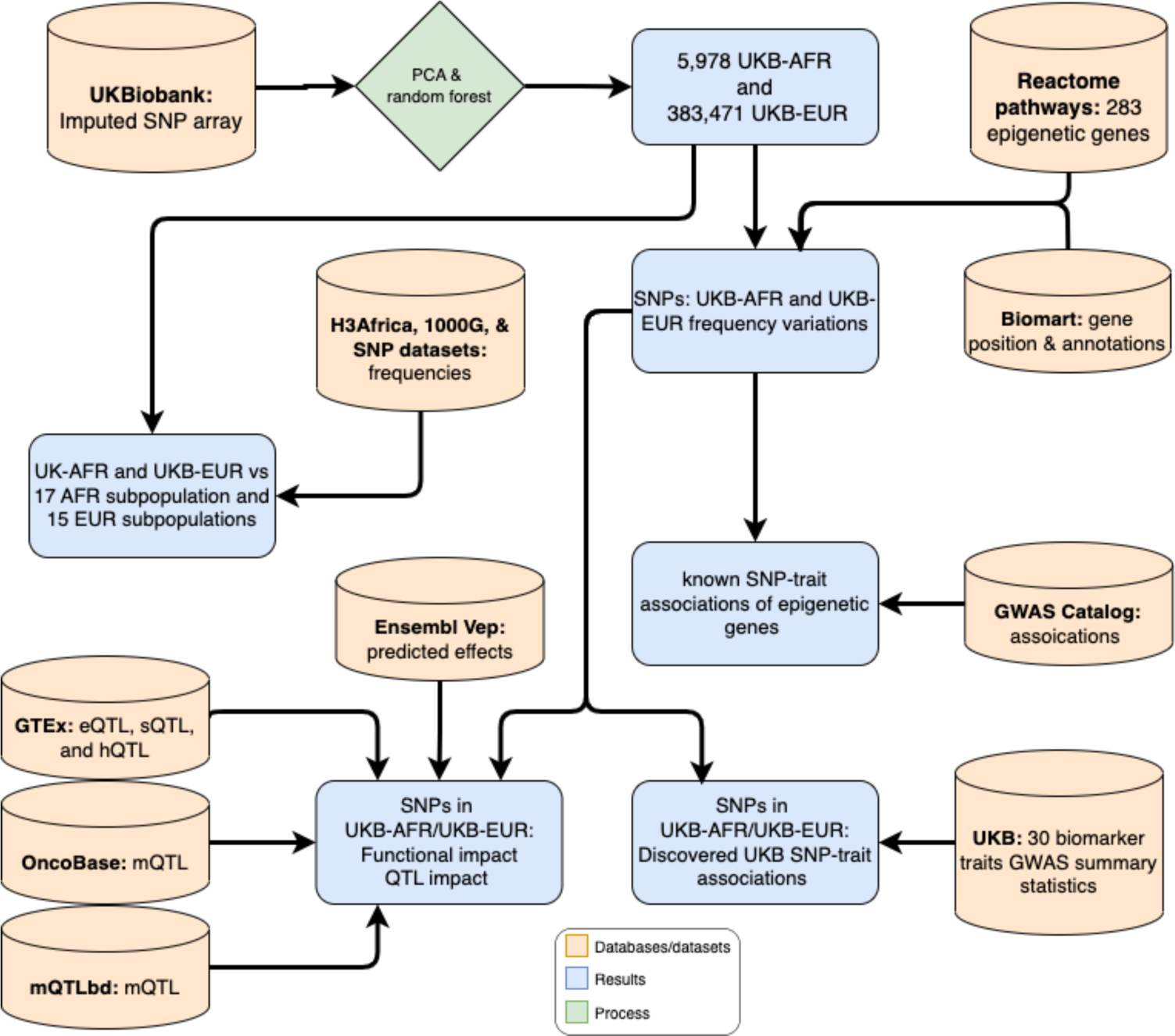
Graphical representation of the overall study method and analysis procedure.

Furthermore, we curated a comprehensive list of 283 epigenetic genes and epigenetic regulator genes across 14 classes from the Reactome pathway knowledgebase ^27,28^ and relevant literature ^29^. This process involved an initial extraction of genes from the Reactome Pathways database, specifically targeting pathways annotated with ‘epigenetic’ descriptors. This step ensured the inclusion of a wide array of genes associated with epigenetic regulation. To further refine our gene list, we consulted supplementary data from Gnad et al. (Supplementary File 8:S5) ^29^, categorising genes into functional groups such as Bromodomain-containing proteins (BROMO), Histone Acetyltransferases (HAT), Histone Deacetylases (HDAC), Histone Demethylases (HDM), Histone Methyltransferases (HMT), and SWI/SNF chromatin remodelling complexes (SWISNF). This dual approach enabled us to assemble a robust and detailed collection of epigenetic genes, pivotal for our study of the impact of genetic variation on epigenetic features and gene expression in AFR populations. In this context, “epigenetic genes” are defined as those involved in epigenetic mechanisms (Figure 9).

### Calculation and comparison of SNP frequencies in AFRs and EURs

We obtained the chromosomal positions of the epigenetic genes from the Ensembl BioMart ^73^ database to map the SNPs to epigenetic genes extending to 500kb up- and down-stream of the gene location. We used the genotyping array data and imputed SNPs from the UKB and returned a subset of the variants in or near an epigenetic gene locus (500kb up- and down-stream of the gene location).

Then, we used the non-reference allele frequency of epigenetic gene variants separately for the 5,978 AFR and 383,471 EUR ancestry individuals previously calculated by the UKB. Briefly, for SNP frequency calculations within the EUR population in the UK Biobank, consider the following hypothetical example: If a specific SNP has a non-reference allele appearing twice among the 383,471 EUR individuals, and since each individual has two alleles at any given position, the total number of alleles in the population is 766,942. The frequency of this SNP’s non-reference allele would be calculated as 2/(2 x 383,471), which simplifies to 1/383,471 = 2.61 x 10^-06^. This calculation method illustrates how the UK Biobank determines SNP frequencies, providing crucial insights into the distribution of genetic variants distribution within specific populations ^74^.

Fisher’s exact test was employed to discern frequency variations in variants between UKB-AFR and UKB-EUR populations within epigenetic gene regions and across all the gene regions, as detailed in Supplementary Figure 19. In addition, we considered variants with Benjamini and Hochberg adjusted p-values < 0.05 as statistically significantly different in their frequencies between UKB-AFR and UKB-EUR.

### Evaluations of variant distribution in AFRs and EURs

To compare the distribution of variants in UKB-AFR and UKB-EUR, we first applied the base ten logarithm to the allele frequencies of each group. Then, we plotted the resulting dataset as frequency histograms separately for UKB-AFR and UKB-EUR (see Figure 2a). Furthermore, to evaluate the magnitude by which alleles vary between UKB-AFR and UKB-AFR, we divided the allele frequencies of UKB-AFR with those of UKB-EUR to obtain the SNP frequency ratios. Then, we calculated the base ten logarithms of the SNP frequency ratios to obtain values centred around 0 (for the difference in allele frequency). Finally, we used absolute SNP frequency ratios to plot the data in Figure 2b so that the values are positive and comparable in one plot.

### Comparative Analysis of Epigenetic Gene SNP Frequencies Among Different Populations

To evaluate the genetic similarity between AFR living in Africa and those in UKB, we obtained whole genome sequence datasets from various AFR and EUR genome sequencing projects, including the H3Africa project ^75^, 1000 Genomes project ^14^, Human Genome Diversity Project, and the 1000 Genomes Project ^14^. We merged SNP frequency data from the UKB with those from the projects mentioned above, resulting in a dataset that encompasses information from two distinct AFR studies spanning several countries (refer to Supplementary Data 1), along with EUR data sourced from the UKB and the 1000 Genomes Project. Notably, since some data are WGS and some from arrays, we only considered overlapping SNPs when merging the datasets, following practices supported by studies such as Truelsen et al, which evaluates the concordance of SNP genotyping across different platforms in human genetics, ensuring the reliability and comparability of our genetic analyses ^31,76,77^.

For each of the genotype datasets from each study, we extracted the variants in or near (±500kb) loci of epigenetic genes using information from Ensembl BioMart ^73^, and calculated the allele frequency of the SNPs. The results were combined in a matrix, from which we removed variants in strong linkage disequilibrium (r^2^ > 0.5). We utilised the LD Link REST API to measure LD between variants, downloading LD data for each variant from LDlink.com ^78,79^. This analysis was conducted using either EUR or AFR as the background population. Variants with an LD threshold (r²) > 0.1 were identified and included in the analysis, ensuring a robust understanding of the linkage disequilibrium patterns. All LD scores and calculations presented in this manuscript were conducted using LD Link.

We employed unsupervised hierarchical clustering to elucidate patterns in epigenetic gene SNP frequencies across different populations ^80^. A matrix representing SNP frequencies in different populations served as the input data. We opted for the Euclidean distance metric to quantify the dissimilarity between data points, owing to its intuitive interpretation and widespread use in genomics, and we selected complete linkage to compute distances between clusters. This approach facilitated the grouping of populations with similar epigenetic gene SNP frequencies, thereby providing insights into their genetic relatedness and diversity.

To further explore the inherent structure of the data, we performed dimensionality reduction using t-distributed Stochastic Neighbour Embedding (t-SNE) ^81^ with a Euclidean distance metric. We subjected the matrix of SNP frequency data to t-SNE and extracted the first two dimensions for visualisation. Plotting these two dimensions on scatter plots enabled the discernment of clustering patterns, where each point represented a population, and the proximity between points indicated similarity in epigenetic gene SNP frequencies.

### Differences in Variant Frequencies Among AFR Populations and the UK Biobank Populations

We then consolidated data from different sources by merging datasets containing SNP frequency data from the various projects for AFR populations as described previously, and the UKB dataset for AFR and EUR populations.

A contingency table was constructed for each SNP for each pair of populations to determine whether the variant frequencies differed significantly between AFR and non-AFR populations, and Fisher’s exact test was applied. The number of significantly differing (p < 0.05) SNPs between each pair of populations was tabulated.

### Functional Impact of Variants in the AFR Population

We started by identifying SNPs with a higher non-reference allele frequency in the UKB-AFR population than in the UKB-EUR population. SNPs with an allele frequency greater than 0.1 in the UKB-AFR population and less than 0.001 in the UKB-EUR population were selected for functional analysis. We used the Ensembl VEP ^32^ via its RESTful API to explore the functional impact of the selected SNPs. The information extracted from the API response included the SNP identifier, the variant’s most severe consequence, and information from the first transcript consequence. The transcript consequence information included the biotype of the transcript and the amino acid change caused by the variant. We combined the information collected from the Ensembl VEP API with the information from the initial set of selected SNPs. This included gene symbols, gene classes, variant identifiers, and allele frequencies in AFR and EUR UKB populations. This dataset provides a comprehensive overview of the selected SNPs with high frequency in the UKB-AFR population and their predicted functional impacts.

### Analysis of GWAS Catalog associations at epigenetic gene loci

We retrieved data from previous GWAS studies from the NHGRI-NBI GWAS Catalog ^82^ and returned traits associated with variants in or near (±500kb) loci of epigenetic genes. To identify variants previously associated with traits in the GWAS Catalog, we extended our analysis to include variants with strong linkage disequilibrium (r^2^ > 0.5) with each other among the 2,444 variants in the GWAS Catalog and with the 223,336 UKB variants. Briefly, for each lead variant (lowest GWAS p-value, i.e., most statistically significant at loci) associated with a trait in the GWAS Catalog and the UKB, we lookup variants associated with the traits to extend the GWAS Catalog to the variants in high linkage disequilibrium (r^2^ ≥ 0.5) with the GWAS hits. If any other variant meets this criterion, we consider it to be the same discovery as the trait’s lead variant. Furthermore, we computed instances where genes of a specific gene class and individual genes were associated with GWAS traits (see Figure 5). Finally, we computed the number of instances in the GWAS Catalog in which each trait is associated with an epigenetic gene.

### Analysis of quantitative trait loci of epigenetic genes

We sought to provide a comprehensive functional annotation and prediction of the regulatory effect of the epigenetic gene variants. First, we collected information on quantitative trait loci (QTLs), which include eQTLs from the GTEx ^65^ and oncoBase ^83^ databases, mQTLs from the mQTLdb ^84^ and oncoBase databases, and hQTLs and sQTLs from GTEx. We returned only mQTLs, sQTLs, eQTLs, and hQTLs that map to regions that encode epigenetic genes, extending to (±500kb) up- and down-stream of the gene location using gene coordinate information from Ensembl BioMart. We integrated the QTL information within variant sets from the UKB to identify QTLs that vary in their frequencies between UKB-AFR and UKB-EUR (see Figures 6 and Supplementary Figure 6). Finally, to identify variants that have multiple QTL, we compared the variant IDs and positions extending to the variants in strong linkage disequilibrium (r^2^ ≥ 0.5) as described above for all QTL (hQTL, eQTL, sQTL, and mQTL) set comparisons.

### Evaluation of the quantitative trait loci effects and trait associations

We integrated the QTL information with data from the GWAS Catalog to identify QTLs associated with various traits and the variation in their frequency between UKB-AFR and UKB-EUR individuals. Furthermore, to explore the relevance of regulatory variation affecting epigenetic gene QTLs in GWAS Catalog traits, we assessed the overlap between QTLs and the GWAS Catalog, extended to include variants in high LD (r^2^ ≥ 0.5, as described above) with the GWAS hits. Then, for each class of QTLs, we computed the number of QTL and non-QTL variants associated with traits and the number of QTL and non-QTL variants not associated with traits. Finally, we applied Fisher’s exact test to computed numbers to determine the extent to which QTLs, compared to non-QTLs, are associated with traits in the GWAS Catalog.

### Integrative analysis of variant frequencies and QTLs in UK Biobank traits

We obtained the GWAS summary statistics computed by the UKB project for 25 blood and urine biomarker traits, including, among others, serum albumin, triglycerides, and cholesterol (see Supplementary Figure 10). The number of AFR and EUR individuals used to compute the GWAS statistics for each biomarker trait is shown in Supplementary Figure 9. Other sources discuss the procedures used to conduct the GWA analyses. ^85^. Briefly, the GWAS was performed for each trait and ancestry group using the Scalable and Accurate Implementation of Generalized Mixed Model Approach ^86^, using a linear or logistic mixed model including a kinship matrix as a random effect and covariates as fixed effects. The covariates included the participant’s age, sex, age multiplied by sex, the square of the age, the square of the age multiplied by the sex, and the first 10 principal components calculated from the genotype datasets. The Manhattan plots were produced in MATLAB using the software described here ^87^. The methods applied for genotyping participants in the UK Biobank are reported elsewhere ^21,88^. Furthermore, the genotyping quality control implemented for the analyses is described at the following link: https://pan.ukbb.broadinstitute.org/docs/qc.

We integrated the UKB GWA significant variants with UKB variant frequency data for EUR and AFR, and the collated QTL information from GTEx, oncoBase, and mQTLdb. The integrated data allowed us to compare the frequency of variants associated with traits only in UKB-EUR, UKB-AFR, and both populations. Furthermore, we used the integrated data to identify QTLs associated with biomarker levels among AFR and EUR in the UKB.

### Correlation of biomarker traits associations in the UK Biobank

We use the UKB GWA information of 25 biomarkers to identify epigenetic gene variants that are shared between the biomarkers by extending to high linkage disequilibrium (r^2^ ≥ 0.5) with the GWA hits in the UKB. Then, we applied unsupervised hierarchical clustering with a Euclidean distance metric using complete linkage to visualise the relationship between shared epigenetic gene variants among the biomarker traits in the UKB. Furthermore, we created a network of biomarker traits. Briefly, for each pair of biomarkers, we defined a link between those two biomarkers if they shared variants (extending to variants in strong LD as described above). In addition, we defined the size of the edges connecting the network nodes (biomarker traits) based on the number of shared associated variants. The resulting connectivity network based on shared variants between the biomarker traits was visualised in the yEd graph visualisation software.

### Statistics and Reproducibility

We used the R programming language, Python, MATLAB 2021a, and Bash to conduct the statistical analysis. We used the Welch test, the Wilcoxon rank-sum test, and the one-way analysis of variance to compare continuous measures among groups. We employed Fisher’s exact test to identify variants with differing frequencies between AFR and EUR populations. All statistical tests were considered significant if the two-sided p-value was < 0.05 for single comparisons. In addition, the Benjamini and Hochberg approach was used to calculate a two-sided q-value (False Discovery Rate) for each group or comparison to correct the multiple hypothesis testing ^89^.

## Data Availability

The raw datasets that support the results presented in this manuscript are available from sources in Table 2. The pre-processed datasets can be accessed via Zenodo (https://doi.org/10.5281/zenodo.12789774)^90^ under the Creative Commons Attribution 4.0.

**Table 2:**
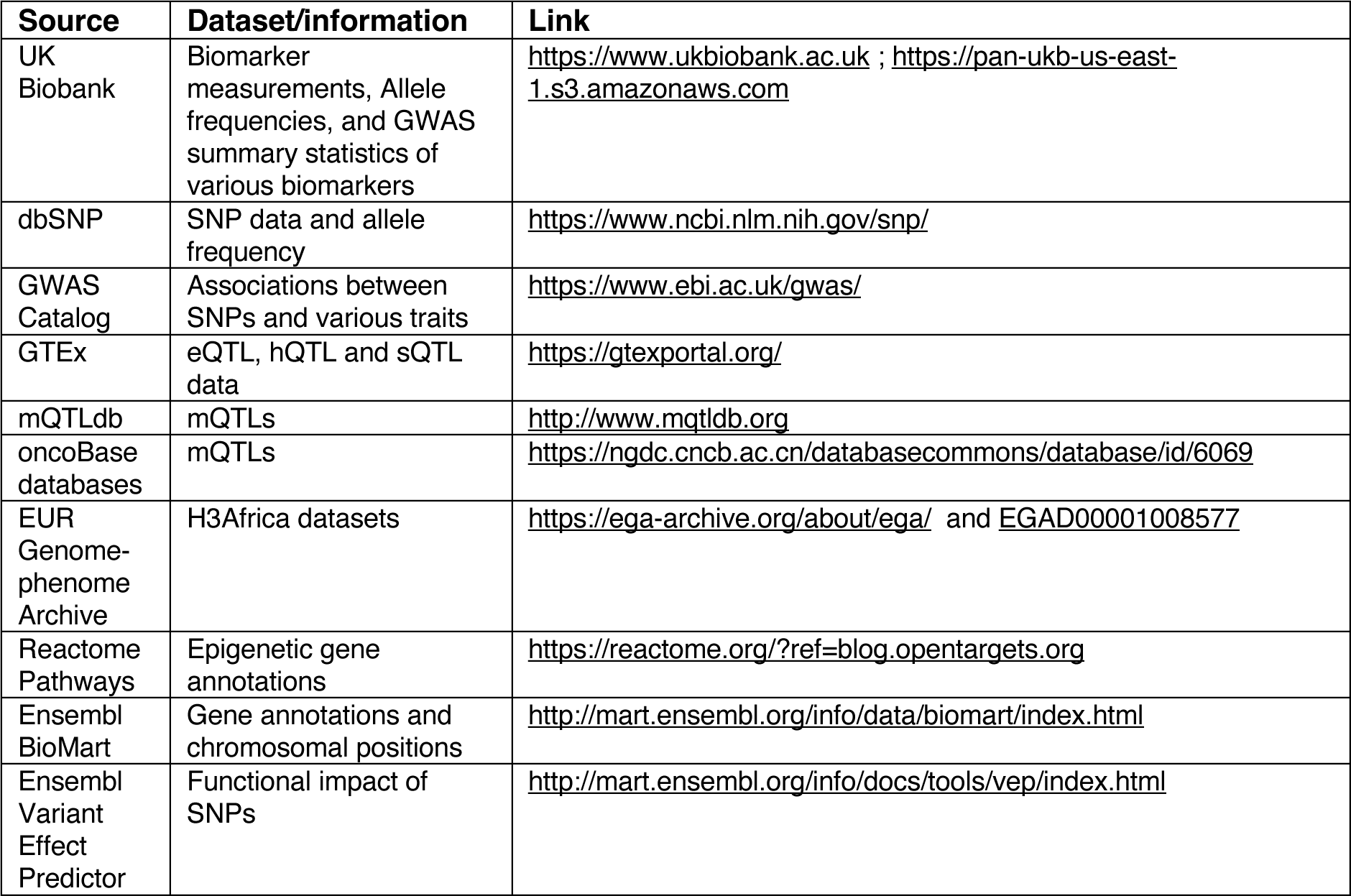
list of resources and datasets.

## Code Availability

Code to reproduce most of the results and plots is available from the following GitHub repository: https://github.com/smsinks/epigenetic-gene-variant-dynamics-analysis.

## Acknowledgements

We would like to acknowledge the contributions of the following individuals for their valuable support and collaboration in this research: Enock Matovu, Busisiwe Mlotshwa, Guida Landoure, Simo Gustaveand, and Martin Simuunza.

This research has been conducted using the UK Biobank Resource under Application Number 53163. The funding for this project was provided by H3ABioNet, supported by the National Institutes of Health Common Fund under grant number U24HG006941. Clement A. Adebamowo and Sally N. Adebamowo were supported by the African Collaborative Center for Microbiome and Genomics Research (ACCME) Grant (1U54HG006947), funds through the Maryland Department of Health’s Cigarette Restitution Fund Program (CH-649-CRF), and the University of Maryland Greenebaum Cancer Center Support Grant (P30CA134274). The content of this publication is solely the authors’ responsibility and does not necessarily represent the official views of the National Institutes of Health.

## Author Contributions

The study was conceptualised by Musalula Sinkala (M.S.), Gaone Retshabile (G.R.), Phelelani T. Mpangase (P.T.M.), Nicola Mulder (N.M.), Salia Bamba (S.B.), Modibo K Goita (M.K.G), Vicky Nembaware (V.N.), Samar S. M. Elsheikh (S.S.M.E.), Jeannine Heckmann (J.H.), Kevin Esoh (K.E.), Mogomotsi Matshaba (M.M.), Ambroise Wonkam (A.W.), Michele Ramsay (M.R.), Clement A. Adebamowo (C.A.A.), Sally N. Adebamowo (S.N.A.), and Ofon Elvis Amih (O.E.A.). The methodology was designed by M.S., G.R., P.T.M., and N.M. Data collection and provision were carried out by M.S., G.R., P.T.M., S.B., M.K.G., V.N., S.S.M.E., J.H., K.E., O.E.A., A.W., M.M., M.R., C.A.A., and S.N.A. Formal analysis of the data was performed by M.S., G.R., S.B., and M.K.G. The manuscript was drafted by M.S., G.R., P.T.M., and N.M. Editing and reviewing of the manuscript were carried out by M.S., G.R., P.T.M., S.B., M.K.G., V.N., S.S.M.E., J.H., K.E., M.M., A.W., M.R., C.A.A., S.N.A., O.E.A., and N.M. M.S produced data visualisations. N.M. supervised the study.

## Ethics approval

The study protocol was approved by The University of Cape Town; Health Sciences Research Ethics Committee IRB00001938.

