## Supplemental Information for "Mapping Epigenetic Gene Variant Dynamics: Comparative Analysis of Frequency, Functional Impact and Trait Associations in African and European Populations"

### 1. Supplementary Figures

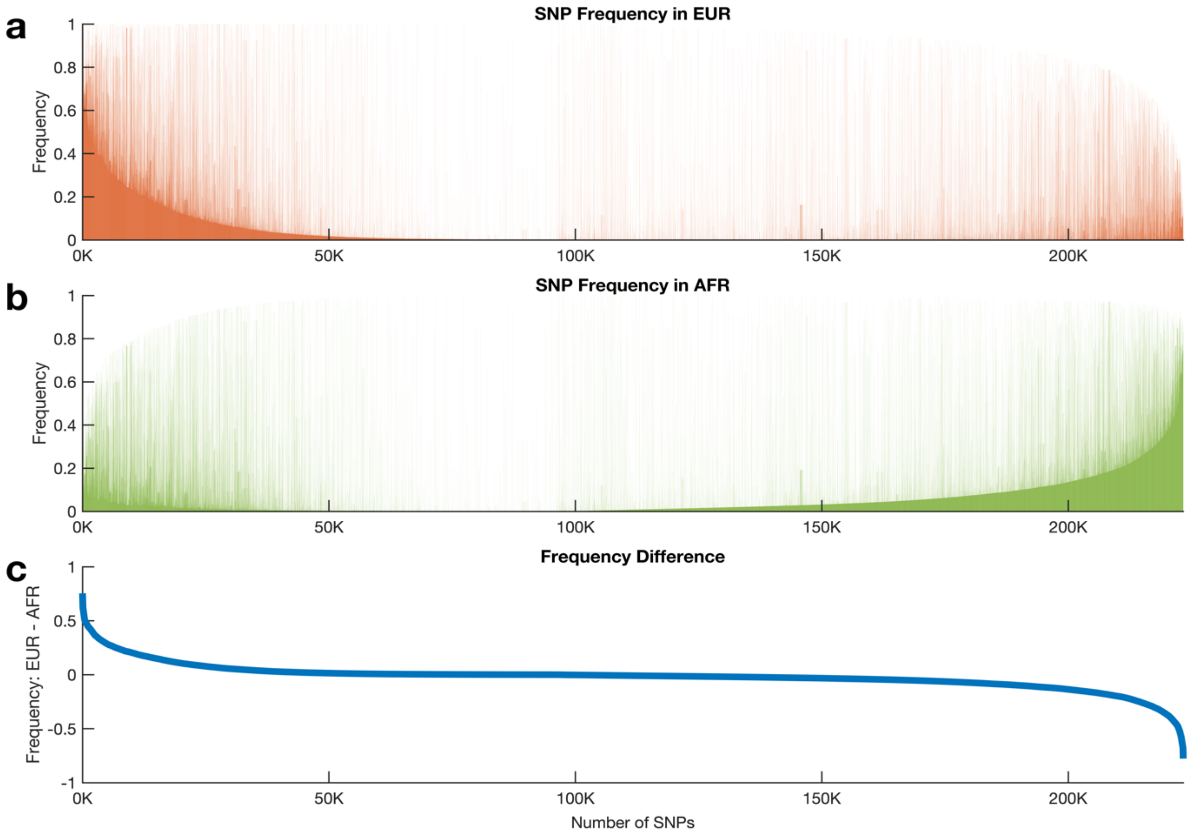


***Supplementary Figure 1:*** *Variation of epigenetic gene SNPs. (a) Frequency of epigenetic gene variants in EUR. (b) Frequency of epigenetic gene variants in AFR. (c) The absolute difference in the frequency of epigenetic gene variation between AFR and EUR. The variants on the x-axis are arranged based on the absolute difference in EUR frequency minus the AFR frequency.*

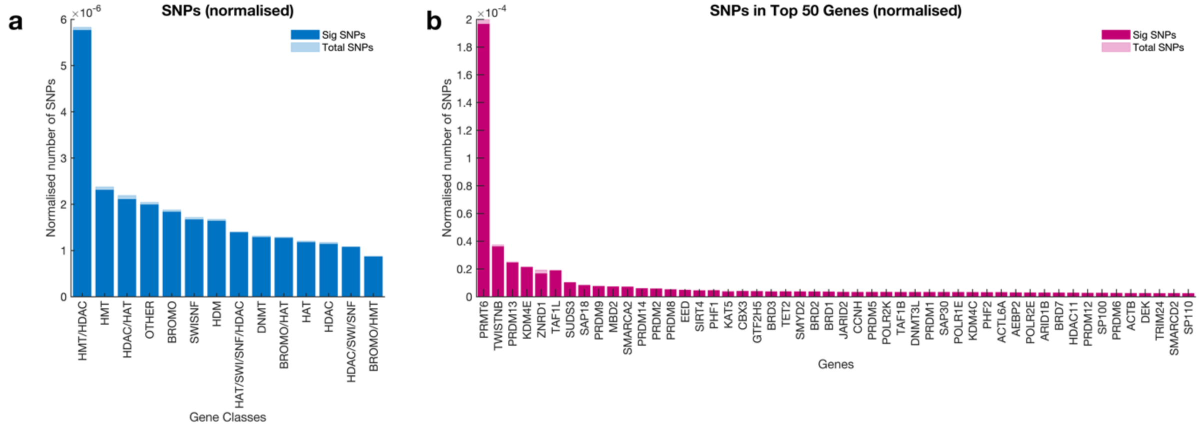


***Supplementary Figure 2:*** *Normalised SNP frequencies of epigenetic genes. (a) The number of variants found in each class of epigenetic genes normalised for the total length of the regions 500kb start and end of the genes in*

*each class. (d) The number of variants in the top 20 epigenetic genes with the most variants normalised for the total length of the gene regions 500kb start and end of the genes.*

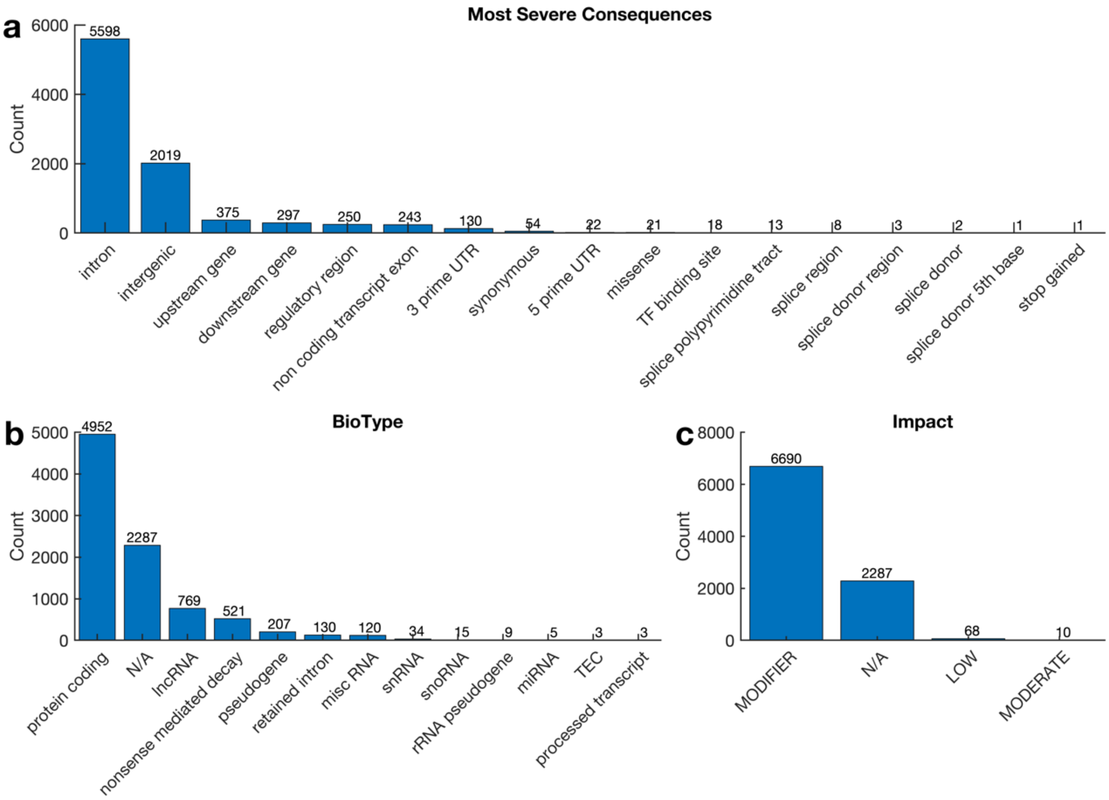


***Supplementary Figure 3:*** *The functional impact of the variant is common in AFR (AF > 0.1) and rare in the EUR (AF < 0.00001) populations. (a) Distribution of the most severe consequences of the common variants in AFR that are rare in the EUR population. (b) the biological types of the genes within which the variants skewered in the AFR population, and (c) their predicted impact on gene or protein function.*

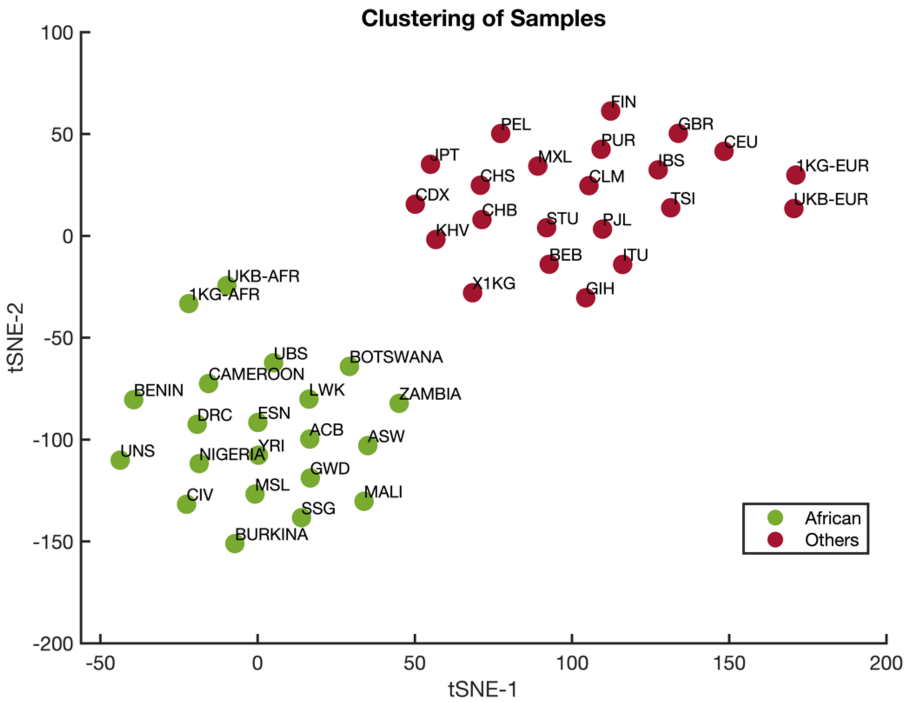


***Supplementary Figure 4:*** *Clustering of populations based on the frequency of all genetic variants located in epigenetic genes. The first two dimensions of the t-SNE reduced datasets are used to show the clustering.*

**Variant Frequency Difference**

| | ACB | ASW | BENIN | BOTSWANA | BURKINA | CAMEROON | CIV | DRC | ESN | GWD | LWK | MALI | MSL | NIGERIA | SSG | UBS | UNS | YRI | ZAMBIA | UKB-AFR | UKB-EUR |
|---|---|---|---|---|---|---|---|---|---|---|---|---|---|---|---|---|---|---|---|---|---|
| ACB | 0 | 747 | 2911 | 4264 | 1258 | 2355 | 771 | 732 | 1185 | 2342 | 2143 | 3853 | 1217 | 2263 | 1279 | 1232 | 4585 | 1079 | 2735 | 14631 | 70440 |
| ASW | 747 | 0 | 4887 | 5218 | 3341 | 3732 | 1716 | 1554 | 2961 | 3327 | 2113 | 4474 | 2836 | 3784 | 2205 | 2798 | 4930 | 2753 | 4293 | 14590 | 49004 |
| BENIN | 2911 | 4887 | 0 | 1507 | 320 | 259 | 374 | 544 | 2485 | 3654 | 6269 | 997 | 2696 | 735 | 597 | 935 | 3620 | 2437 | 980 | 14849 | 62233 |
| BOTSWANA | 4264 | 5218 | 1507 | 0 | 1413 | 856 | 1343 | 692 | 4115 | 5323 | 4360 | 2726 | 4088 | 1621 | 1803 | 771 | 3641 | 4467 | 902 | 17550 | 61005 |
| BURKINA | 1258 | 3341 | 320 | 1413 | 0 | 316 | 276 | 480 | 1369 | 1808 | 3431 | 751 | 1168 | 407 | 399 | 305 | 2696 | 1468 | 585 | 11602 | 44604 |
| CAMEROON | 2355 | 3732 | 259 | 856 | 316 | 0 | 446 | 312 | 1947 | 3004 | 3932 | 612 | 2356 | 292 | 446 | 458 | 2837 | 2202 | 557 | 13977 | 63778 |
| CIV | 771 | 1716 | 374 | 1343 | 276 | 446 | 0 | 225 | 907 | 1270 | 2600 | 888 | 522 | 520 | 169 | 681 | 2681 | 890 | 838 | 12089 | 57022 |
| DRC | 732 | 1554 | 544 | 692 | 480 | 312 | 225 | 0 | 503 | 1750 | 1125 | 966 | 829 | 381 | 264 | 261 | 1719 | 963 | 306 | 11693 | 52392 |
| ESN | 1185 | 2961 | 2485 | 4115 | 1369 | 1947 | 907 | 503 | 0 | 2439 | 2223 | 4967 | 1385 | 2050 | 1338 | 1620 | 5374 | 215 | 3305 | 16317 | 66015 |
| GWD | 2342 | 3327 | 3654 | 5323 | 1808 | 3004 | 1270 | 1750 | 2439 | 0 | 5559 | 3657 | 1097 | 2853 | 620 | 2752 | 6185 | 2028 | 4216 | 20192 | 65050 |
| LWK | 2143 | 2113 | 6269 | 4360 | 3431 | 3932 | 2600 | 1125 | 2223 | 5559 | 0 | 7118 | 3566 | 4007 | 3183 | 963 | 4007 | 2543 | 4025 | 20497 | 62551 |
| MALI | 3853 | 4474 | 997 | 2726 | 751 | 612 | 888 | 966 | 4967 | 3657 | 7118 | 0 | 3609 | 1342 | 847 | 1218 | 3408 | 4686 | 1835 | 16470 | 56354 |
| MSL | 1217 | 2836 | 2696 | 4088 | 1168 | 2356 | 522 | 829 | 1385 | 1097 | 3566 | 3609 | 0 | 2317 | 488 | 1358 | 5019 | 1412 | 3813 | 17093 | 62717 |
| NIGERIA | 2263 | 3784 | 735 | 1621 | 407 | 292 | 520 | 381 | 2050 | 2853 | 4007 | 1342 | 2317 | 0 | 837 | 729 | 1833 | 2061 | 1106 | 13634 | 53157 |
| SSG | 1279 | 2205 | 597 | 1803 | 399 | 446 | 169 | 264 | 1338 | 620 | 3183 | 847 | 488 | 837 | 0 | 648 | 2843 | 1250 | 1156 | 13882 | 60891 |
| UBS | 1232 | 2798 | 935 | 771 | 305 | 458 | 681 | 261 | 1620 | 2752 | 963 | 1218 | 1358 | 729 | 648 | 0 | 1216 | 1757 | 600 | 12437 | 43664 |
| UNS | 4585 | 4930 | 3620 | 3641 | 2696 | 2837 | 2681 | 1719 | 5374 | 6185 | 4007 | 3408 | 5019 | 1833 | 2843 | 1216 | 0 | 5238 | 3609 | 18876 | 53161 |
| YRI | 1079 | 2753 | 2437 | 4467 | 1468 | 2202 | 890 | 963 | 215 | 2028 | 2543 | 4686 | 1412 | 2061 | 1250 | 1757 | 5238 | 0 | 3564 | 17002 | 72822 |
| ZAMBIA | 2735 | 4293 | 980 | 902 | 585 | 557 | 838 | 306 | 3305 | 4216 | 4025 | 1835 | 3813 | 1106 | 1156 | 600 | 3609 | 3564 | 0 | 15501 | 57822 |
| UKB-AFR | 14631 | 14590 | 14849 | 17550 | 11602 | 13977 | 12089 | 11693 | 16317 | 20192 | 20497 | 16470 | 17093 | 13634 | 13882 | 12437 | 18876 | 17002 | 15501 | 0 | 102854 |
| UKB-EUR | 70440 | 49004 | 62233 | 61005 | 44604 | 63778 | 57022 | 52392 | 66015 | 65050 | 62551 | 56354 | 62717 | 53157 | 60891 | 43664 | 53161 | 72822 | 57822 | 102854 | 0 |

Colour scale: $\times 10^4$ — 0, 1, 2, 3, 4, 5, 6, 7, 8, 9, 10

***Supplementary Figure 5:*** *Absolute difference of variants that demonstrate a statistically significant difference between each population as determined using the Fisher exact test. The UKB-AFR and UKB-EUR represent the UK Biobank populations. See Supplementary Data 1 for the number of sample genotypes for each population.*

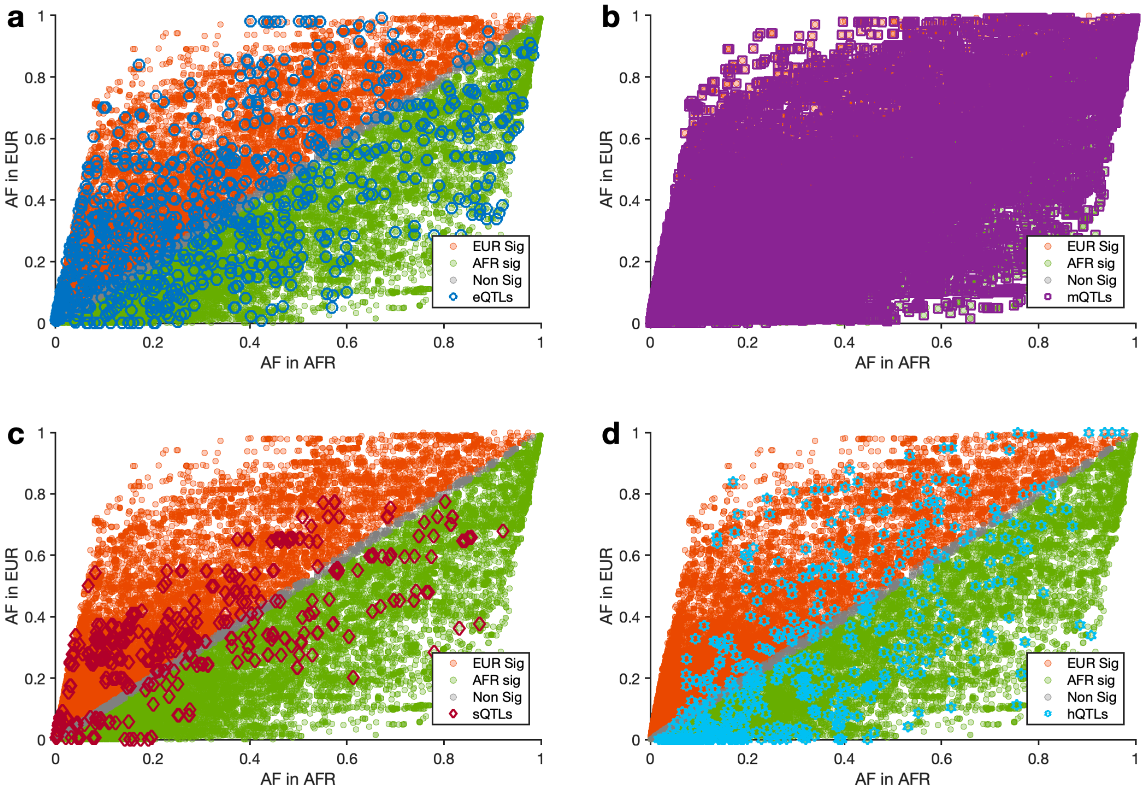


***Supplementary Figure 6:*** *The frequency of all quantitative trait loci of epigenetic genes in AFR and EUR. The frequency of (a) eQTL, (b) mQTL, (c) sQTL, and (d) hQTL variants among AFR and EUR. For all four figure panels, the orange points are for variants frequent in EUR, green for variants significantly more frequent in AFR, and grey for variants whose frequencies are similar in AFR and EUR. Within each panel, the marked points are eQTLs (blue), mQTLs (purple), sQTLs (magenta), and hQTLs (cyan).*

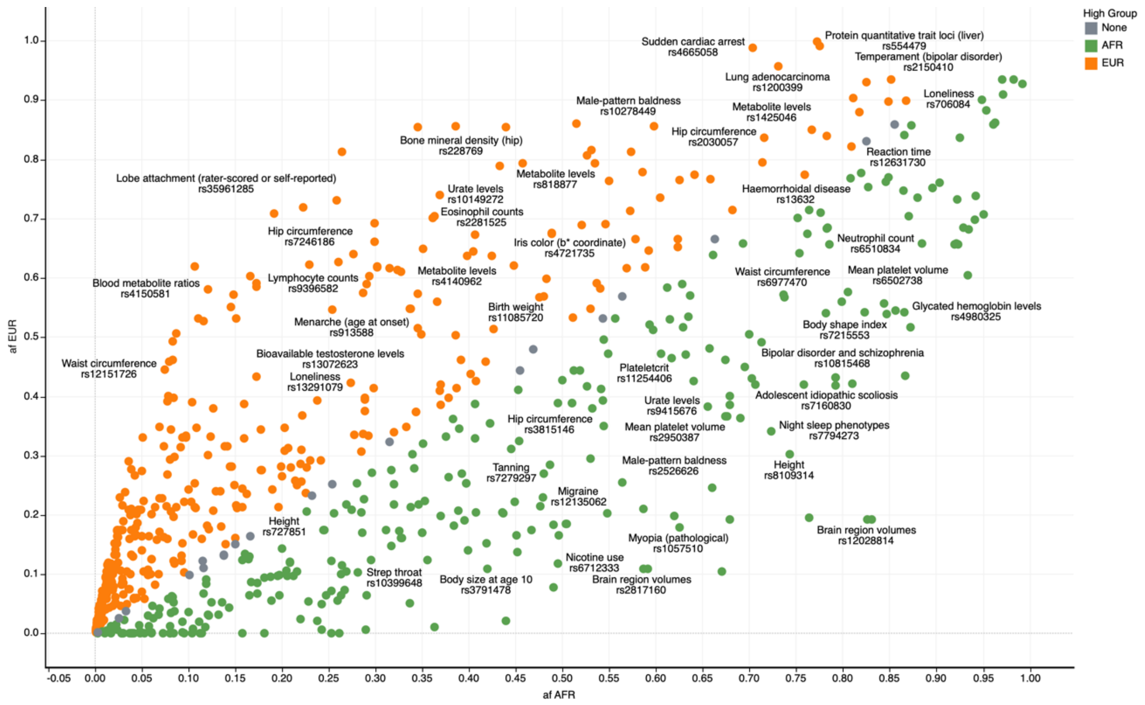


***Supplementary Figure 7:*** *Frequency of eQTLs associated with traits in the GWAS Catalog. The orange points indicate variants that are significantly more frequent in EUR, green for variants that are significantly more frequent in AFR, and grey for variants whose frequencies are similar in AFR and EUR. Some labels are omitted to improve the readability of the figure.* An interactive visualisation of Supplementary Figure 7 can be found here.

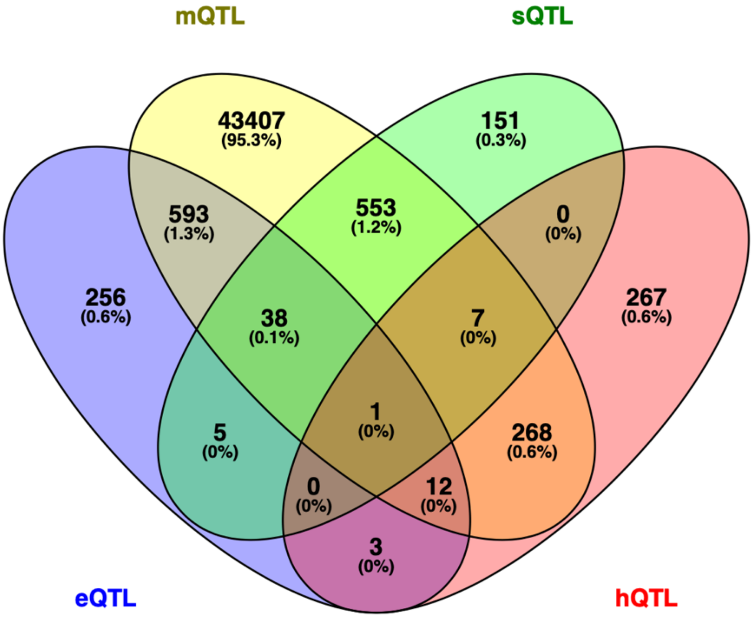


***Supplementary Figure 8:*** *Venn diagram of epigenetic gene variants that are quantitative trait loci.*

| GWAS Trait | mQTL | eQTL | sQTL | hQTL | High Group<br>AFR | EUR | None | Grand Total |
|---|---|---|---|---|---|---|---|---|
| No | False | False | False | False | 105,424 | 68,530 | 3,973 | 177,927 |
| | | | | True | 223 | 37 | 6 | 266 |
| | | | True | False | 45 | 92 | 14 | 151 |
| | | True | False | False | 111 | 135 | 6 | 252 |
| | | | | True | 3 | | | 3 |
| | | | True | False | 3 | 2 | | 5 |
| | True | False | False | False | 16,863 | 24,361 | 1,735 | 42,959 |
| | | | | True | 146 | 110 | 10 | 266 |
| | | | True | False | 139 | 407 | 6 | 552 |
| | | | | True | 2 | 4 | | 6 |
| | | True | False | False | 202 | 351 | 20 | 573 |
| | | | | True | 7 | 5 | | 12 |
| | | | True | False | 13 | 24 | 1 | 38 |
| | | | | True | | 1 | | 1 |
| Yes | False | False | False | False | 112 | 161 | 5 | 278 |
| | | | | True | 1 | | | 1 |
| | | True | False | False | 2 | 2 | | 4 |
| | True | False | False | False | 163 | 271 | 14 | 448 |
| | | | | True | 2 | | | 2 |
| | | | True | False | | 1 | | 1 |
| | | | | True | | 1 | | 1 |
| | | True | False | False | 8 | 11 | 1 | 20 |
| Grand Total | | | | | 123,469 | 94,506 | 5,791 | 223,766 |

High Group
- AFR
- EUR
- None
- Grand Total

***Supplementary Figure 9:*** *Distribution of epigenetic gene variants across quantitative trait loci. The number of epigenetic gene variants is broken down by ancestry groups (High Group), with a statistically significantly higher frequency of variants versus the GWAS Catalog trait. Also, the variants are broken down into mQTL, eQTL, sQTL, and hQTL. The colour shows details about the ancestral group with the significantly higher frequency of the SNPs.* An interactive visualisation of Supplementary Figure 9 can be found here.

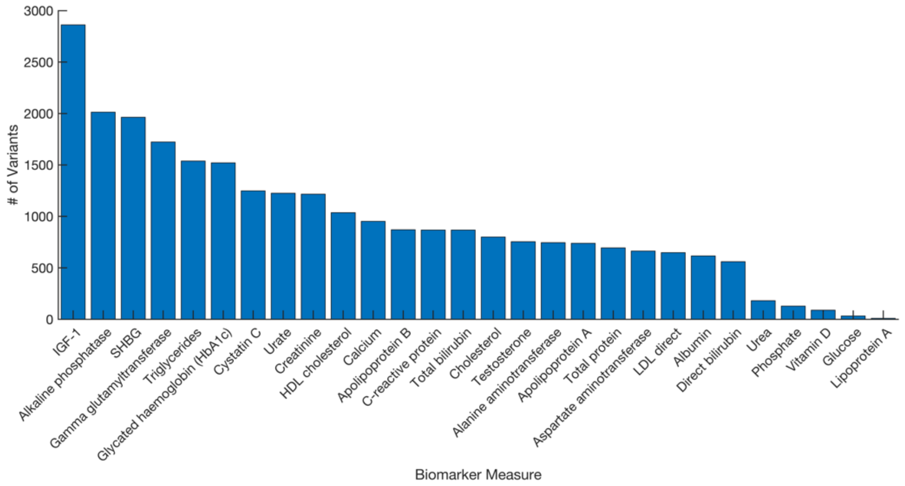


***Supplementary Figure 10:*** *Distribution of epigenetic gene variants significantly associated with 28 blood and urine biomarkers in the UK Biobank.*

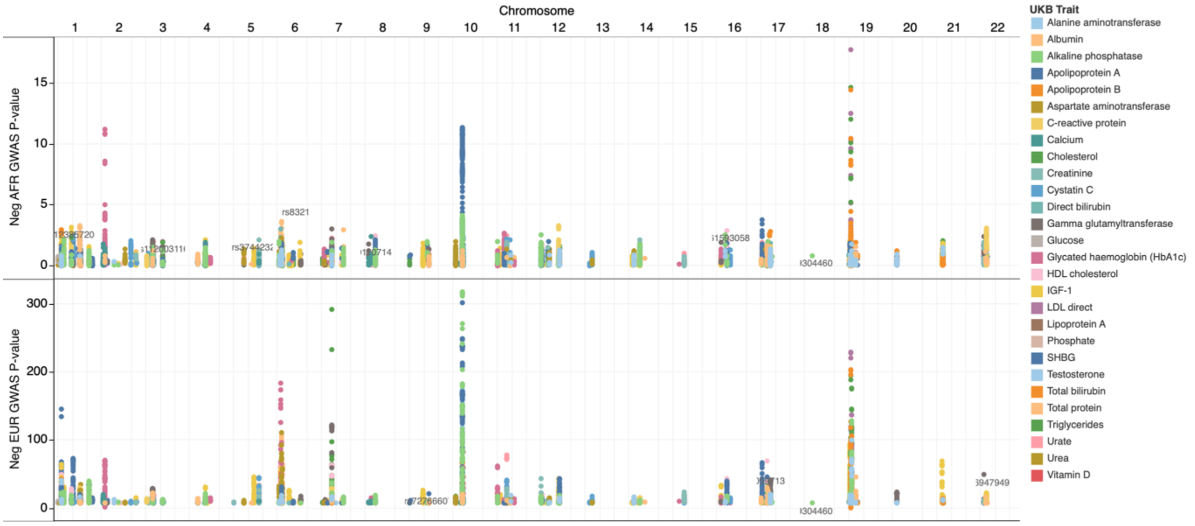


***Supplementary Figure 11:*** *Manhattan plot of epigenetic gene loci associated with 28 biomarker traits in the UK biobank. The figure is broken down by ancestry group, i.e., AFR and EUR. The colours show the biomarker traits associated with the SNPs at various loci.* An interactive visualisation of this figure can be found [here].

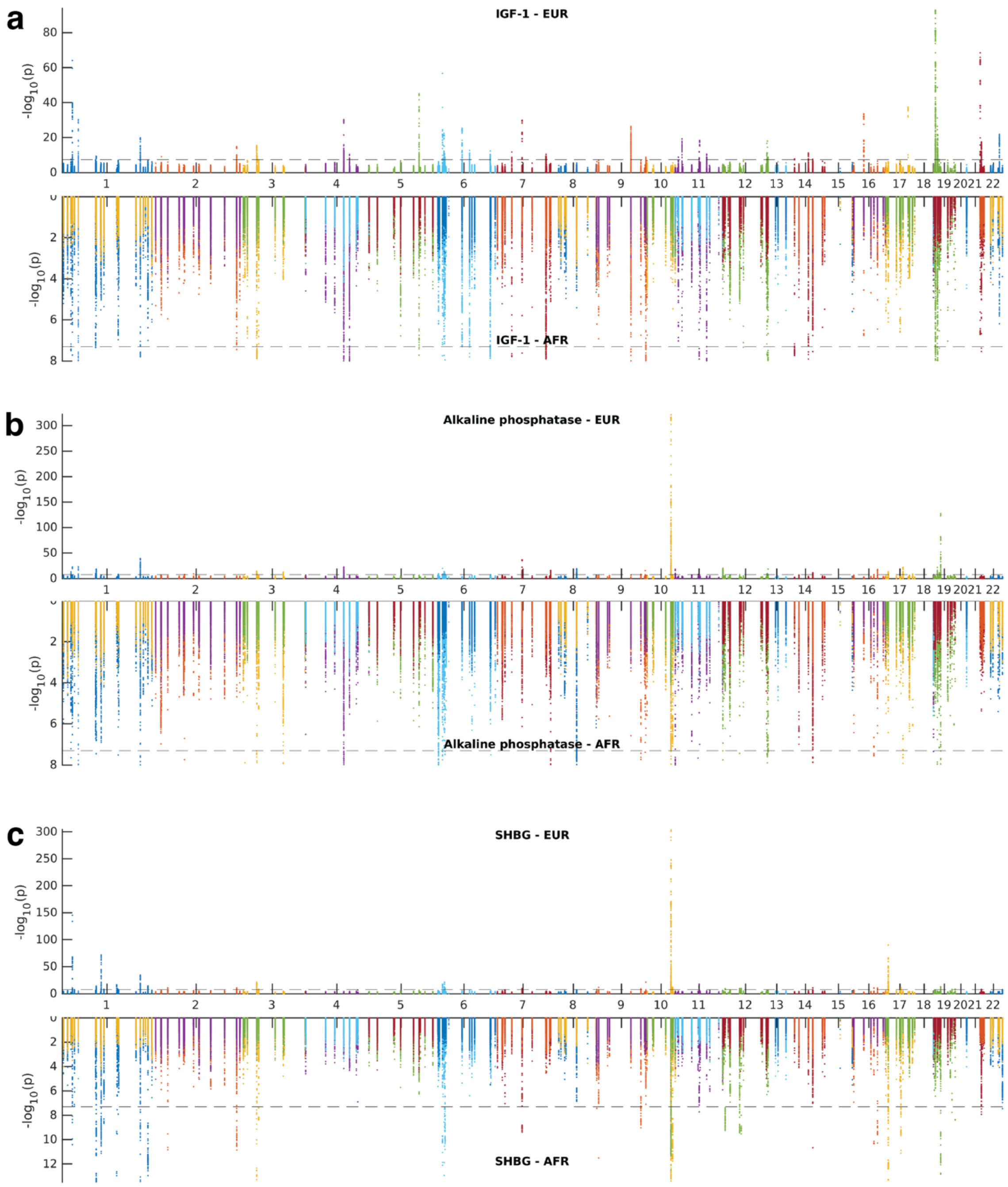


***Supplementary Figure 12:*** *The Manhattan plots for SNPs associated with **(a)** IGF-1 in Europeans and Africans, **(b)** alkaline phosphatase in Europeans and Africans, and **(c)** steroid hormone-binding globulin in Europeans and Africans.*

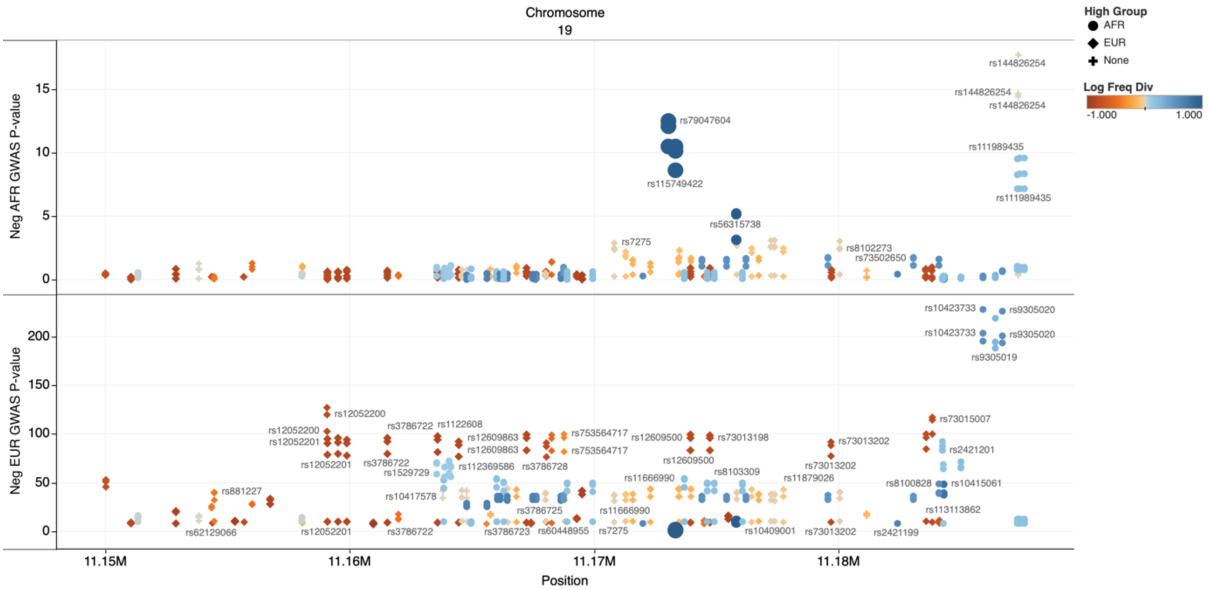


***Supplementary Figure 13:*** Chromosomal regional plots at the SMARCA4 loci. The plot shows the SMARCA4 chromosomal positions vs base-10 negative logarithm GWAS-p-values for biomarker traits associated with AFR and EUR. The chromosomal positions are filtered, ranging from 11,150 KB to 26,845 KB of chromosome 19. The colours show details about the population with a significantly higher frequency of the variants, with bluer colours indicating a higher frequency in AFR and redder colours indicating a higher frequency in EUR. The marker sizes show base 10 absolute logarithms of SNP frequency in AFR vs EUR. The shapes show details about the group in which variants are significantly more frequent. The marks are labelled by the variant SNPdb IDs. An interactive visualisation of Supplementary Figure 13 can be found [here].

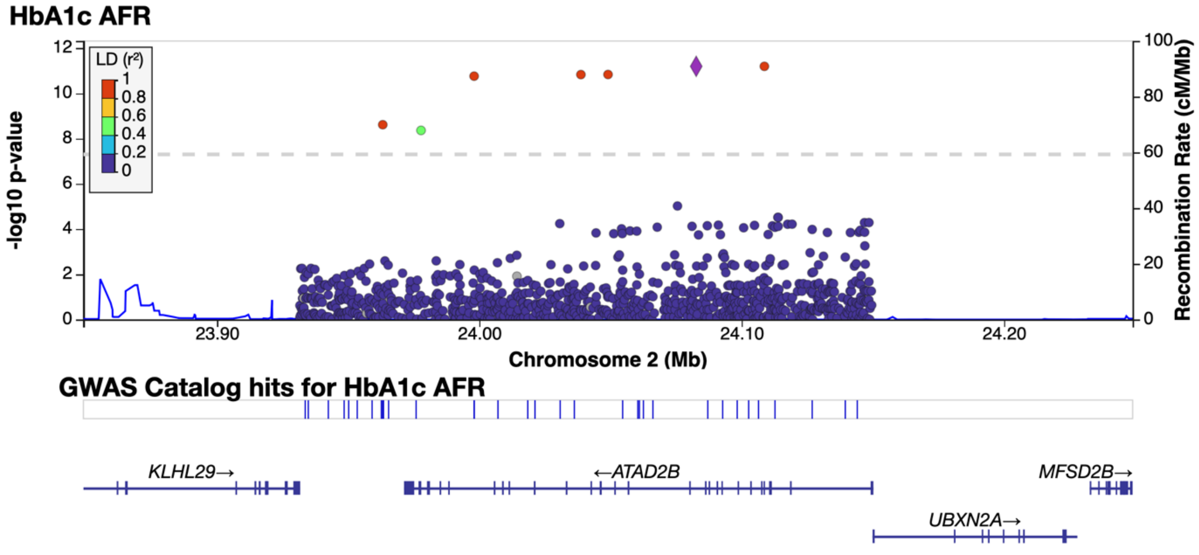


***Supplementary Figure 3:*** *Regional association plots for variants associated with Hb1Ac in AFR. At these loci, the lead SNPs rs114686503. The genes within the chromosomal loci are shown in the lower panel. The blue line indicates the recombination rate. The filled circles show the position of the SNPs along the region on the x-axis and the negative logarithm of the association p-value on the y-axis. The lead SNP (rs114686503) is shown in purple. The SNPs within the locus are coloured based on the linkage disequilibrium correlation value ($r^2$) with the lead SNP based on the African HapMap haplotype from the 1000 Genome project. Also, see Figure 7b.*

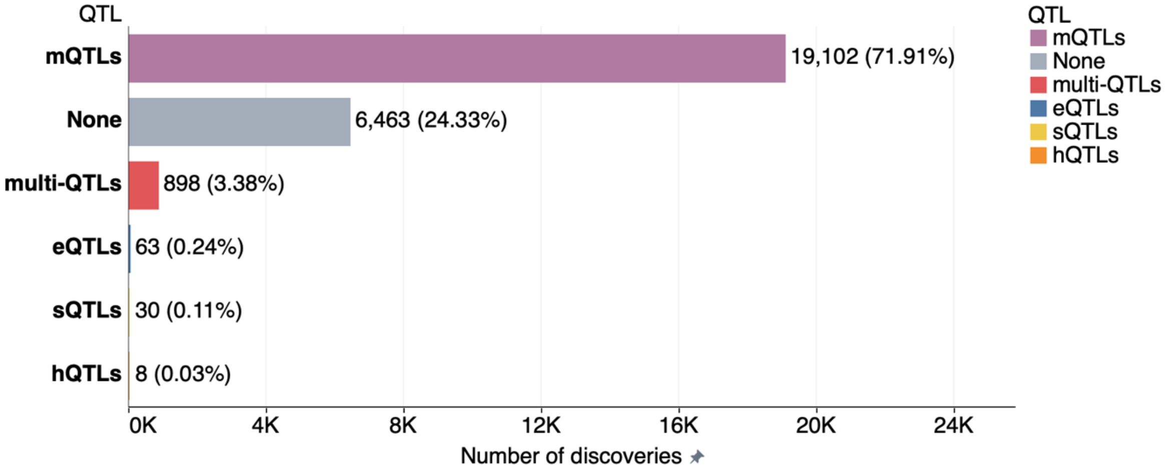


***Supplementary Figure 15:*** *The functional impact of epigenetic variants associated with 28 biomarker traits in the UK Biobank.*

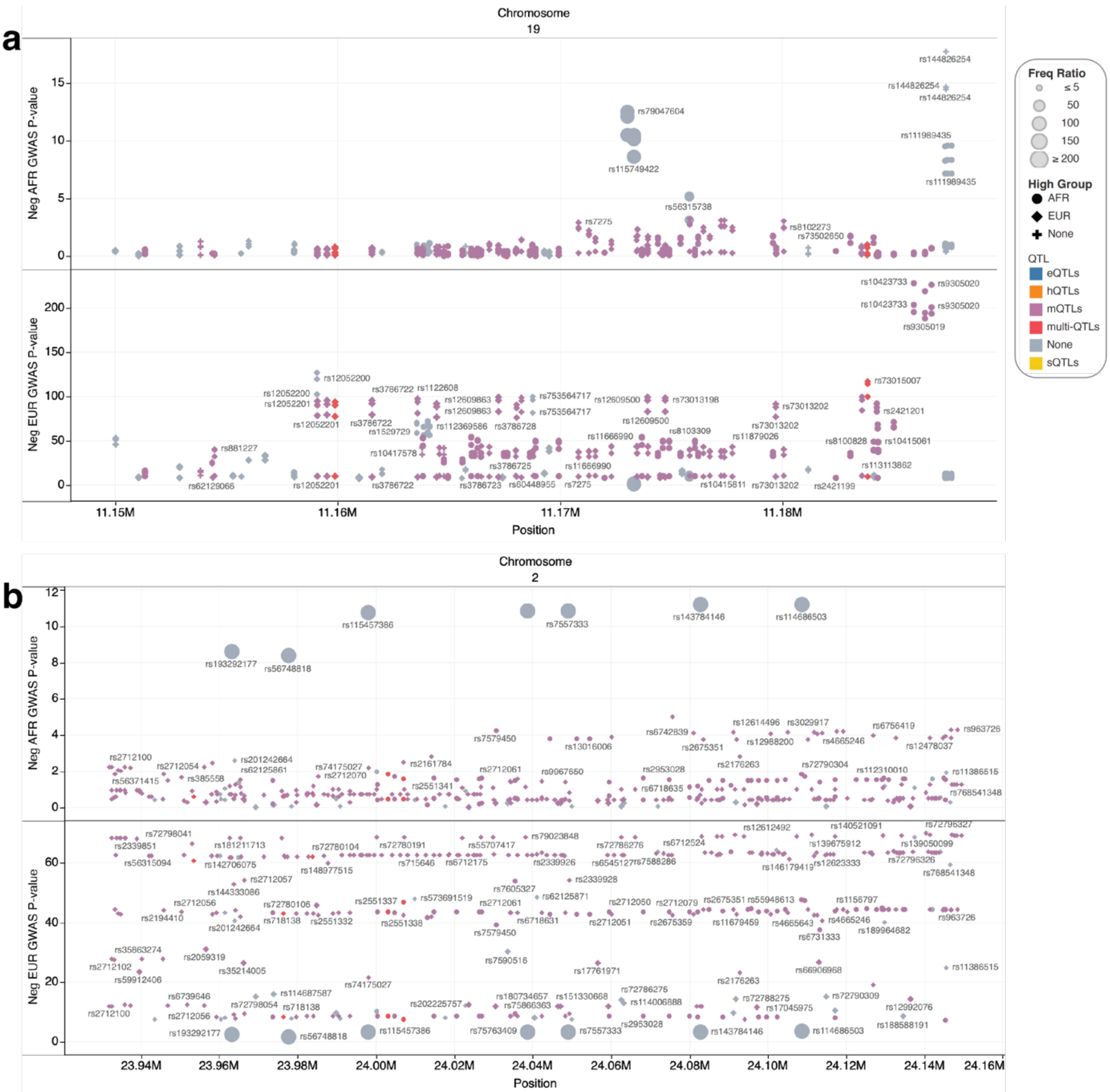


***Supplementary Figure 16:*** *Chromosomal regional plots at the SMARCA4 and ATAD2B gene loci. (a) SMARCA4 chromosomal positions vs base-10 negative logarithm GWAS-p-values for biomarker traits associated*

*with AFR and EUR. The chromosomal positions are filtered, ranging from 11,150 KB to 26,845 KB of chromosome 19. (b) ATAD2B chromosomal positions vs base-10 negative logarithm GWAS-p-values for biomarker traits associated with AFR and EUR. The chromosomal position filter ranges from 23,000 KB to 24,300 KB. The colours show details about quantitative trait loci, whereas the marker sizes show the base 10 absolute logarithms of SNP frequency in AFR vs EUR. The shapes show details about the group in which variants are significantly more frequent. The variant SNPdb IDs are used to label the marks.* An interactive visualisation of Supplementary Figure 16 can be found [here](here).

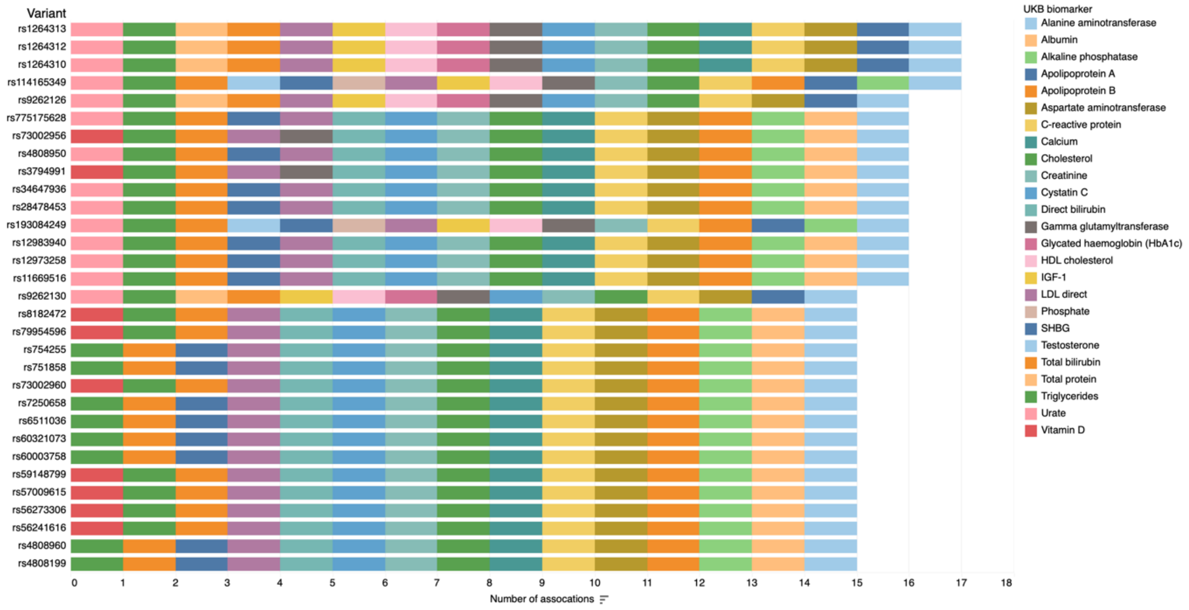


***Supplementary Figure 17:** Epigenetic gene variants associated with multiple biomarker traits in the UK Biobank. The colours show details about the biomarker trait shown in the legends.* An interactive visualisation of Supplementary Figure 17 can be found [here](here).

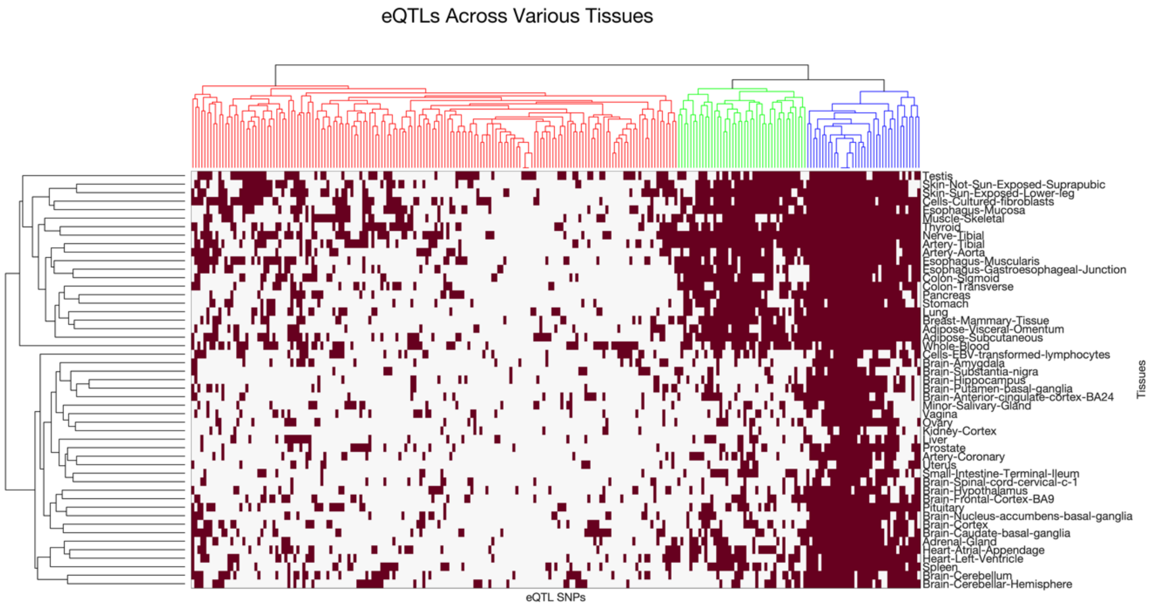


***Supplementary Figure 18:** Clustergram of eQTLs Across Various Tissues Based on GTEx Data. The distribution of eQTLs across a range of human tissues. Each column in the heatmap corresponds to a specific eQTL SNP, while each row represents a different tissue type. The presence of an eQTL in each tissue is indicated by a red mark, whereas its absence is represented by white. The hierarchical clustering on the top (columns) groups eQTLs that share similar patterns across tissues, and the side dendrogram (rows) clusters tissues based on their eQTL profiles. The clustering reveals patterns of epigenetic gene variants specific to certain tissues and those shared across multiple tissues, providing insights into the genetic architecture of gene expression regulation across the human body.*

### 2. Supplementary Data

**Supplementary Data 1:** an Excel workbook that provides detailed annotations and analyses related to the impact of common SNPs found in AFR populations on various genomic elements and functions. The workbook is structured into several sheets, each focusing on different aspects of SNP data:

1. Impact of AFR Common SNPs: This sheet details the effects that common SNPs in African populations have on genomic functions or structures. It likely includes data on how these SNPs influence gene expression, protein function, or other genomic features.
2. Biotype of AFR Common SNPs: This sheet categorises the common SNPs by their genomic locations or functions, such as whether they are located in coding regions, non-coding regions, or regulatory elements.
3. Functional Impact: This sheet provides information on the functional consequences of the SNPs. It may include predictions or observations regarding the impact of these SNPs on protein structure, gene regulation, or disease associations.
4. Genotype Data Populations: This sheet includes data on the frequency of each SNP within different populations, possibly focusing on comparisons between African populations and others or highlighting the diversity within AFR populations.

**Supplementary Data 2:** An Excel workbook that provides comprehensive details on the GWAS traits, genes, and genetic variants reported in the GWAS Catalog, with a focus on their prevalence in African (AFR) and European (EUR) populations. The workbook is organized into several sheets, each highlighting different aspects of the GWAS data:

1. All epigenetic traits: This sheet lists all the epigenetic traits and their associated genetic variants.It includes the prevalence of these variants in both AFR and EUR populations.
2. GWAS AFR Trait: This sheet focuses on trait associated with genetic variants in African populations. It provides a detailed count and description of the variants linked to each tr.
3. GWAS EUR Traits: This sheet details Traits associated with genetic variants in European populations. It includes a count and description of the variants linked to each Trait.
4. All Traits Count: This sheet provides a comprehensive count of all Traits linked to genetic variants, combining data from both AFR and EUR populations. It offers a comparative analysis of Trait prevalence between the two populations.
5. Top 10 Traits: This sheet highlights the top 10 Traits with the highest number of associated genetic variants. It provides a detailed comparison of the prevalence of these Traits in AFR and EUR populations.
6. AFR Trait Count: This sheet focuses on the count of Traits specifically in the African population.It provides a breakdown of the number of genetic variants associated with each Trait in AFR.

7. EUR Trait Count: This sheet focuses on the count of Traits specifically in the European population. It provides a breakdown of the number of genetic variants associated with each Trait in EUR.
8. Gene Class Count: This sheet categorizes the genes based on their classes and provides a count of genetic variants within each class. It includes a comparative analysis of the gene classes between AFR and EUR populations.

**Supplementary Data 3:** an Excel workbook with comprehensive details on genetic variants associated with various UK Biobank biomarkers. Each entry in the dataset is detailed with multiple attributes:

1. HugoSymbol: The gene symbol associated with the SNP.
2. GeneClass: Gene classifications such as HAT, SWISNF, etc.
3. Variant: SNP identifier.
4. Chrom: Chromosome number where the SNP is located.
5. Position: Position of the SNP on the chromosome.
6. ref: Reference allele.
7. alt: Alternative allele.
8. af_AFR and af_EUR: Allele frequencies in African and European populations, respectively.
9. highGroup: The population group where the allele frequency is higher.
10. FDR: False discovery rate.
11. eqtl, mqtl, hqtl, sqtl: Boolean values indicating whether the SNP is an eQTL, mQTL, hQTL, or sQTL, respectively.
12. pval_AFR and pval_EUR: P-values in African and European populations.
13. UKB_trait_type: The type of trait linked with the variant in the UK Biobank.
14. UKB_description: A description of the UK Biobank trait.
15. rep_pvalue: Replication p-value.
16. rep_Variant: Variant used for replication studies.
17. gwasSigGroup: The group significance in GWAS.
18. freqDiv, freqDiff, freqDivProper: Statistical measures of frequency divergence between populations.
19. QTL: The type of QTL the variant represents.

**Supplementary Data 4** provides a comprehensive analysis of biomarker interrelationships within a dataset. This dataset details the count of shared genetic associations between various pairs of biomarkers. Each row in the dataset represents a pair of biomarkers and the number of genetic loci they share, highlighting the extent of their genetic overlap and potential shared biological pathways.

### 3. Supplementary Notes

**Supplementary Note 1:** Differential Frequency and Impact of Epigenetic Variants on Biomarker Traits in African and European Populations

In this study, we have comprehensively analysed the frequencies, functional impacts, and trait associations of epigenetic gene variants across African and

European populations. Our research unveils many findings, highlighting the complexity and richness of epigenetic variation and its consequences on biomarker traits. Given the extensive data generated, the task of statically visualising our findings, particularly in the context of comparing significance across different linkage disequilibrium blocks, assessing frequency differences, evaluating the functional impact of variants, and understanding trait associations, presented significant challenges.

To surmount these obstacles, we utilised interactive visual analytics facilitated by Tableau. This method allowed for an unbiased and dynamic examination of the epigenetic gene variants, offering insights into their differing impacts on African and European populations. Our approach includes the use of interactive Manhattan plots, regional plots with impact annotations, annotations for GWAS p-values and biomarker traits, and figures comparing frequencies, among others, all of which are accessible via provided links:

1. https://public.tableau.com/app/profile/musalula.sinkala7788/viz/EpigeneticVariantTraitsandFrequencies/AllGWASTraits. This visualisation has SNP frequency comparison plots and function impact annotation of SNPs. It also includes an interactive version of several figures and supplementary figures in our manuscript.

2. https://public.tableau.com/app/profile/musalula.sinkala7788/viz/UKBTraitsMappedtoVariantsinAFRandEUR/eQTLtraitlegend. This visualisation has Manhattan plots, regional plots, and quantitative trait loci plots. It also includes an interactive version of several figures and supplementary figures in our manuscript.
3. 

The interactive visualisations at these links contain multiple tabs of plots that allow for:
1. An all-encompassing visualisation and comparison of significant variants associated with various biomarker traits and quantitative trait loci across the 22 chromosomes in both populations.
2. Precise navigation to specific chromosomal locations to highlight differences between African and European variants frequencies, among other aspects.
3. Advanced interrogation of variants within or adjacent to specific genes facilitated by gene-centric filters in the visualisation tools.
4. Custom filtering of variants based on p-value thresholds, chromosome, gene, variant, or genomic position enables detailed comparative analyses and reveals differences in genetic architecture between the two groups.

These interactive analytical tools have significantly enhanced our understanding of the genetic variants located within epigenetic genes, their frequency differences between individuals of African and European ancestry, their functional impacts, and their associations with traits.

**Supplementary Note 2:** Variation in Frequencies of All Genetic Variants in the UKB

The analysis was expanded to include all variants to provide a comprehensive understanding of genetic variation in epigenetic genes. This broader analysis compared allele frequencies of variants across the genome between AFR and EUR populations in the UKB. Findings indicated that 51.23% (52.3% for genes vs. 51.1% for epigenetic genes) of variant alleles were more prevalent in the AFR cohort, and 36.72% (36.7% for genes vs. 37.3% for epigenetic genes) were more common in the EUR cohort, with 10.96% of variants showing no significant difference. This detailed examination, encompassing 15,143,796 variants higher in AFR and 10,631,111 higher in EUR, confirms the initial observations of reduced variation within AFR sub-populations compared to differences between AFR and EUR populations (see Supplementary Figures 1a to 1c). Notably, 89.03% of all variants demonstrated significant differences between these populations, emphasising the nuanced genetic diversity within and between groups.

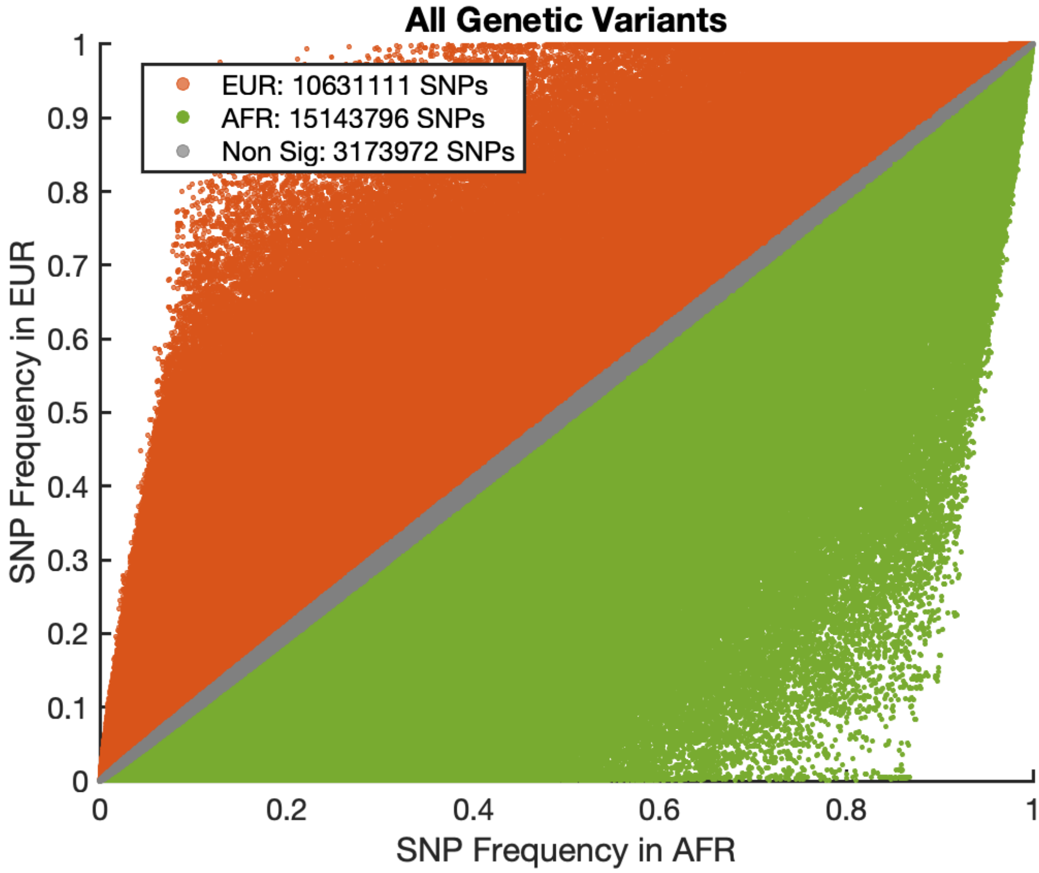


**Supplementary Figure 19:** *Disparity in SNP frequencies between African and European populations, encompassing the complete dataset of variants in the UK Biobank. The scatter plot illustrates the minor allele frequencies for all evaluated genetic variants, juxtaposing AFR and EUR populations. SNPs are depicted as dots, with colour coding indicative of their prevalence: red for a higher frequency in EURs (10,631,111 SNPs), green for a higher frequency in AFRs (15,143,796 SNPs), and orange for those with non-significantly different frequencies (3,173,972 SNPs). The identity line highlights instances of equal allele frequencies, underscoring the significant genetic variation observed between these populations.*
